# Scalable, high quality, whole genome sequencing from archived, newborn, dried blood spots

**DOI:** 10.1101/2022.07.27.22278102

**Authors:** Yan Ding, Mallory Owen, Jennie Le, Sergey Batalov, Kevin Chau, Yong Hyun Kwon, Lucita Van Der Kraan, Zaira Bezares-Orin, Zhanyang Zhu, Narayanan Veeraraghavan, Shareef Nahas, Matthew Bainbridge, Joe Gleeson, Rebecca J. Baer, Gretchen Bandoli, Christina Chambers, Stephen F. Kingsmore

## Abstract

Universal newborn screening (NBS) is an incredibly successful public health intervention. Archived dried bloodspots (DBS) collected for NBS represent a rich resource for population genomic studies. To fully harness this resource, DBS must yield high-quality genomic DNA (gDNA) for whole genome sequencing (WGS). In this pilot study, we hypothesized that gDNA of sufficient quality and quantity for WGS could be extracted from archived DBS up to 20 years old without PCR (Polymerase Chain Reaction) amplification. We describe simple methods for gDNA extraction and WGS library preparation from several types of DBS. We tested these methods in DBS from 25 individuals who had previously undergone diagnostic, clinical WGS and 29 randomly selected DBS cards collected for NBS from the California State Biobank. While gDNA from DBS had significantly less yield than from EDTA blood from the same individuals, it was of sufficient quality and quantity for WGS without PCR. All samples DBS yielded WGS that met quality control metrics for high-confidence variant calling. Twenty-eight variants of various types that had been reported clinically in 19 samples were recapitulated in WGS from DBS. There were no significant effects of age or paper type on WGS quality. Archived DBS appear to be a suitable sample type for WGS in population genomic studies.

## INTRODUCTION

Newborn dried blood spots (DBS) are used worldwide to screen for childhood genetic diseases with effective treatments. Over the past 50 years, universal newborn screening (NBS) has proven an incredibly successful public health intervention for reducing morbidity and mortality due to certain selected conditions.^1–8^ Archived DBS represent the largest repository of human genetic material in existence.^2,5,9,10^ In the United States, approximately 4 million newborns are screened each year, and some states store DBS from these infants. The California Biobank Program represents the combined biospecimen and data resources of the California Genetic Disease Screening Program and the California Birth Defects Monitoring Program. These programs began tracking birth defects in 1983, and currently screen approximately 400,000 newborns each year for treatable, infant onset diseases. Currently, the California Recommended Uniform Screening Panel includes 33 primary disorders and 51 secondary disorders.^2,9,11^ Newborn screening is currently undertaken at California Department of Health Services screening laboratories, and consists of specific assays for each set of disorders. Currently, DNA sequencing is not part of the primary screen, and is only undertaken as a confirmatory test in some states after a primary screen returns an abnormal result.

DBS represent an invaluable resource for research aimed at elucidating the underlying etiology of human diseases, including birth defects, metabolic disorders, and congenital heart defects, and may be particularly suitable for newly available methods for whole genome sequencing (WGS).^1,6,12,13^ Additionally, for infants who die shortly after birth, DBS may be resourced for investigation of the molecular cause of death.^14–16^ DBS have numerous advantages over whole blood samples, including ease of transport, potential for storage at room temperature, cost effectiveness and feasibility of long-term storage without degradation of DNA.^10,17–20^ Despite the success of newborn screening, a significant number of treatable genetic diseases go undiagnosed at birth.^14–16^ The cost and time required for WGS has rapidly decreased over the past decade.^21–24^ In the future, newborn screening for genetic diseases with effective treatments could be greatly expanded by WGS of DBS.

Several issues have been previously identified when using archived DBS for WGS. It has been suggested that the age of the DBS, storage conditions, and filter paper type may impact the yield and quality of the sequencing data.^10,17–20,25–27^ Previous studies have shown mixed results when examining these potential sources of variability. For example, Hollegaard et al. found a significant effect of storage length on quality of extracted DNA in samples dating back to 1981.^17^ However, the sequencing data still demonstrated an average call rate of 97% for single nucleotide variants (SNVs).^12^ This result was replicated in a recent study by Sok et al.^27^ Most studies have examined whole-exome sequencing (WES) from DBS, not WGS. More recently, Bassaganyas et al. reported using DNA isolated from archived DBS to perform WES and WGS with limited polymerase chain reaction (PCR) cycling post-ligation and concluded that the DBS were a satisfactory source of high quality DNA.^20^ Of note, none of these studies evaluated structural variant calls. Finally, previous studies specifically assessing WGS from DBS have included small sample sizes and the gDNA yield has been low.

WGS using archived DBS samples from publicly held biorepositories presents the potential to investigate genetic disease at a population level. In this pilot study, we hypothesized that using the newest techniques for WGS without PCR amplification would result in high-quality sequence irrespective of the DBS age or filter paper type. We describe simple, scalable methods for WGS on laboratory created DBS and randomly selected, de-identified DBS from the California Biobank Program.

## RESULTS

### Quality and quantity of genomic DNA extracted from manufactured DBS

We manufactured 63 DBS from EDTA-blood samples remnants from 25 individuals who had previously received diagnostic PCR-free WGS on Illumina Novaseq 6000 instruments (Table S1). In 19 individuals, at least one variant had been reported clinically (Table 1). DBS were made with two types of filter paper that are widely used for NBS (FTA, ThermoFisher and PC, GE Healthcare; Table 1). Genomic DNA (gDNA) was extracted without noticeable degradation from both types of DBS using two different sample preparation methods (Illumina PCR-free genomic library preparation [Illumina] and KAPA HyperPlus [QIAGEN]; Figure 1; Table S1). The QIAGEN isolation method generated more gDNA per extraction than Illumina (average 868 ng and 349 ng, respectively, p<0.01, minimum 369 ng and 165 ng, respectively; Table S2). gDNA from DBS had a slightly smaller molecular weight than that isolated from the corresponding EDTA-blood but was acceptable for library construction (Figure 1). The A260/A280 ratio of gDNA generated with QIAGEN was higher than that of Illumina (average 1.72 and 1.57, respectively, p<0.01; Table S2). In comparison, 350 µL EDTA-blood from the same individuals yielded an average of 6,884 ng gDNA with an A260/A280 ratio of 1.81 (Table S2). The filter paper type (FTA or PC DBS) did not affect the concentration of gDNA extracted nor the A260/A280 ratio (p>0.05).

**Table 1:** Concordance of variants reported clinically in diagnostic WGS of 25 blood samples and WGS from 63 manufactured DBS. Abbreviations: P, pathogenic; VUS, variant of uncertain significance; LP, likely pathogenic; del, deletion; dup, duplication. *Confirmed diagnosis using standard annotation, variant alignment and analysis pipelines as detailed in Methods.

| Subject ID | Relationship to Proband | Variants previously reported following diagnostic WGS | Variant classification | FTA DBS WGS | PC DBS WGS | Dx Confirmed* |
| --- | --- | --- | --- | --- | --- | --- |
| 1 | Proband | *FOXP3*, c.1010G>A, p.Arg337Gln | P | 2 | 3 | Y |
| 2 | Proband | None | None | 1 | 2 |  |
| 3 | Proband | Chr22:23961084-24401339del | P | 1 | 2 | Y |
| 4 | Father | None | None | 2 | 2 |  |
| 5 | Sibling | *FLNA*, c.2410G>A, p.Val804Ile | VUS | 1 | 2 | Y |
| 6 | Proband | *FLNA*, c.2410G>A p.Val804Ile; Chr2:112658998-112854380dup | VUS | 1 | 2 | Y |
| 7 | Proband | *DCLRE1C*, c.406G>A, p.Asp136Asn; Chr10:14983601-15065700del | LP, P | 1 | 2 | Y |
| 8 | Proband | *RYR2*, c.12290A>G, p.Asn4097Ser; *ANK2*, c.8404G>C, p.Asp2802His | VUS | 1 | 1 | Y |
| 9 | Father | *RYR2*, c.12290A>G, p.Asn4097Ser; *ANK2*, c.8404G>C, p.Asp2802His | VUS | 1 | 1 | Y |
| 10 | Proband | *SMN1* Chr5:70247540-70247820x0 del; *SMN2* Chr5:69372123-69372400dup | P | 2 | 3 | Y |
| 11 | Proband | *TUBB3*, c.1228G>A, p.Glu410Lys; Chr22:18873501-21466000del | P | 3 | 3 | Y |
| 12 | Proband | *ROBO1*, c.107G>T, p.Arg36Met; *ROBO1*, c.4610G>A, p.Gly1537Glu | VUS | 1 | 2 | Y |
| 13 | Proband | *PROKR2*, c.563C>T, p.Ser188Leu; Chr14:59001701-61049600dup | VUS | 2 | 2 | Y |
| 14 | Proband | *HSD17B4*, c.1619A>G, p.His540Arg | VUS | 1 | 2 | Y |
| 15 | Proband | None | None | 1 | 1 |  |
| 16 | Proband | None | None |  | 1 |  |
| 17 | Proband | None | None |  | 1 |  |
| 18 | Proband | *EPG5*, c.2066del, p.Leu689Ter | P | 1 | 1 | Y |
| 19 | Proband | *EPG5*, c.2066del, p.Leu689Ter | P | 1 | 1 | Y |
| 20 | Proband | None | None | 1 |  |  |
| 21 | Proband | Chr15:23512201-28700800del | P | 1 |  | Y |
| 22 | Proband | Chr10:81634801-89151100 del | P | 1 |  | Y |
| 23 | Proband | Chr2:21240919-21244369del | P | 1 |  | Y |
| 24 | Proband | ChrX:1422154-1423912del | P | 1 |  | Y |
| 25 | Proband | *SMN1* Chr5:70247540-70247820x0del; *SMN2* Chr5:69372122-69372400x2 dup | P | 1 |  | Y |

**Table 2.**
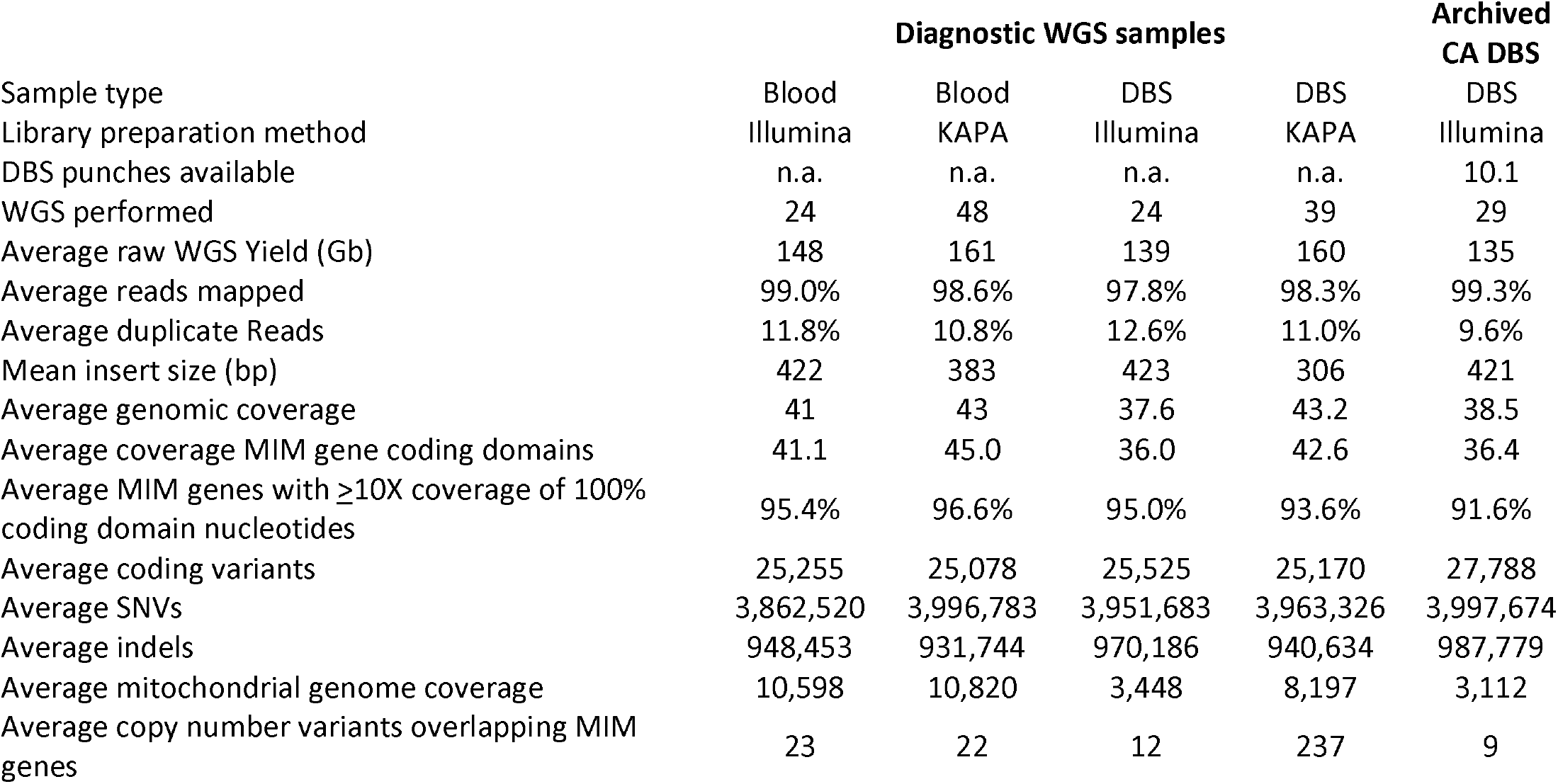
Quality of diagnostic WGS of 25 blood samples (controls) compared with WGS from 63 DBS made from those blood samples and 29 archived California newborn DBS. Two library preparation methods were used (Illumina PCR-free genomic library preparation and KAPA HyperPlus). Abbreviation: MIM, Mendelian Inheritance in Man; CA, California.

**Figure 1.**
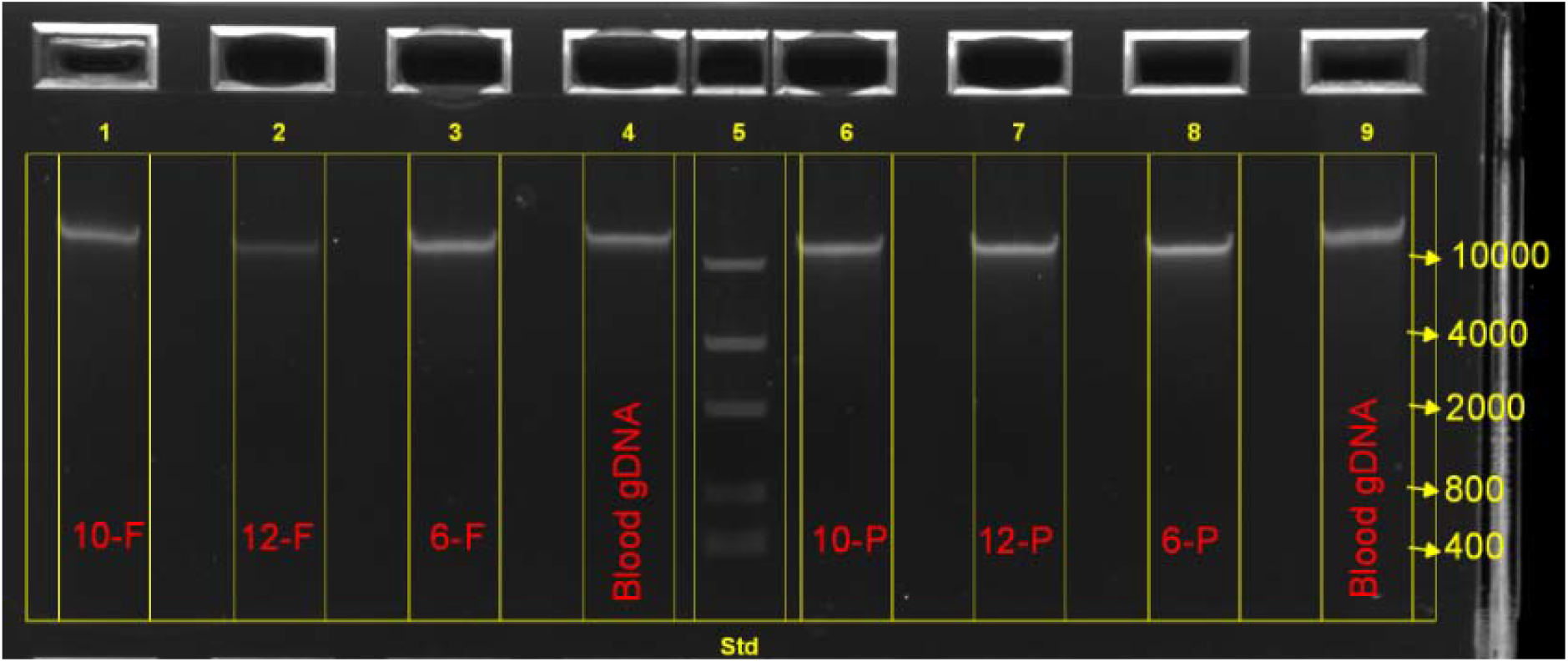
Image of electrophoresis of genomic DNA from DBS samples 10-FTA, 12-FTA, 6-FTA, 10-PC, 12-PC, 6-PC and blood in a 0.8% agarose gel. Molecular weight standards are shown (nucleotides). A single high-molecular weight band is observed, with no apparent DNA degradation.

### Titration of quantity of genomic DNA extracted from DBS punches

A significant issue in using archived DBS is the number of punches available. Often, DBS have undergone prior extractions for NBS or research purposes, and therefore the number of punches available is decreased. We performed titration experiments with six DBS to evaluate the minimal number of 3 mm diameter punches required for WGS. The median gDNA isolated from 1, 3, 5, 7, and 10 punches was 53 ng, 123 ng, 216 ng, 421 ng, and 707 ng, respectively (Figure 2a). Thus, six DBS punches were required to isolate at least 200 ng gDNA per extraction. The DNA yield decreased with the length of storage of manufactured DBS (Figure 2a). The A260/A280 ratio did not vary with age of DBS (Figure 2b).

**Figure 2:**
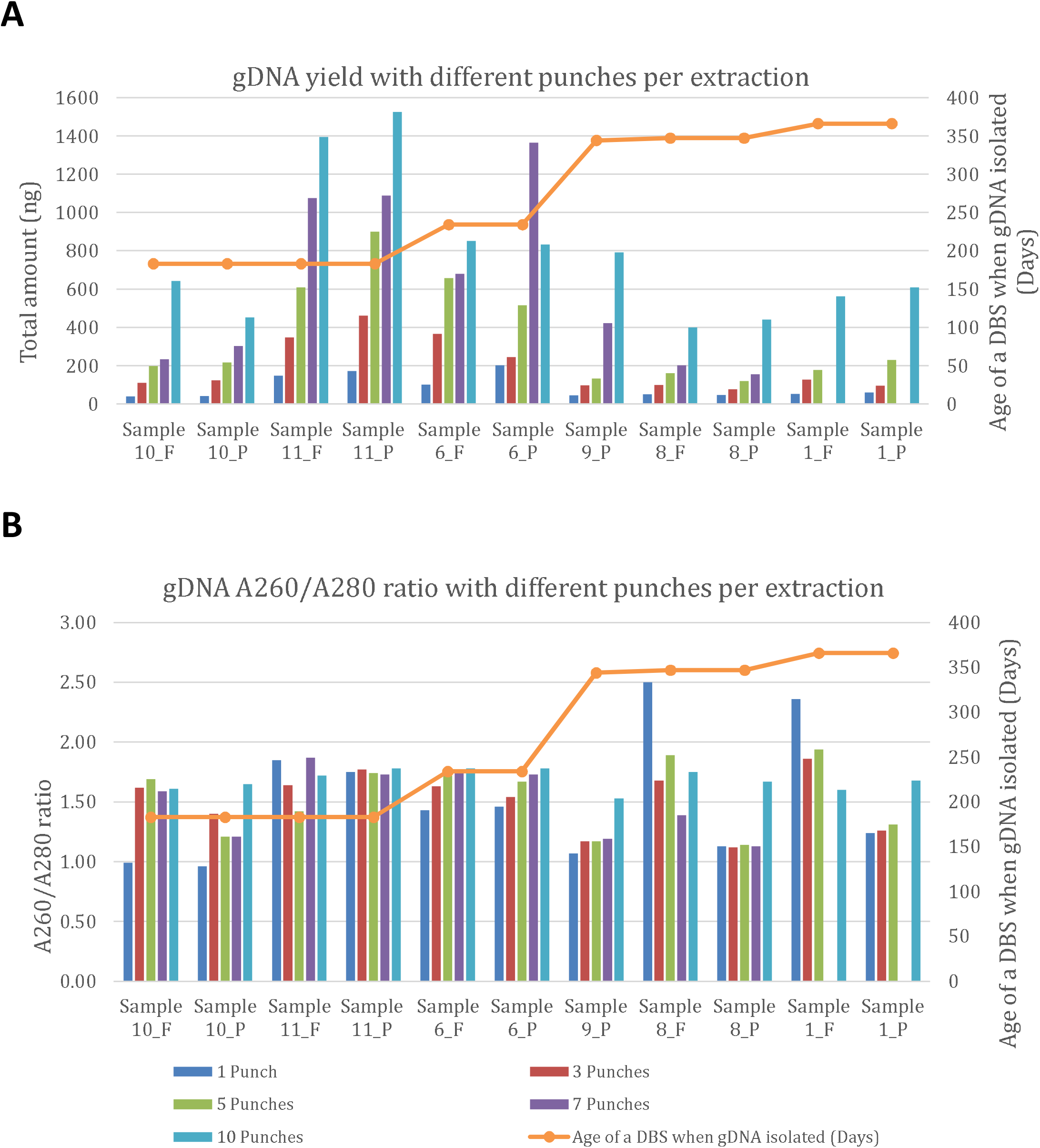
Relationship of number of DBS punches and genomic DNA yield (a) and A260/A280 ratio (b) for 11 DBS. Samples were 1, 6, 8, 9, 10 and 11. DBS were made with FTA and PC filter papers, with the exception of sample 9 (PC only).

### Quality of sequencing libraries generated from DBS-derived genomic DNA

All 63 gDNA samples yielded sequencing libraries without requirement for PCR. There were no library failures. The range of library yield was 3–18 nM, and mean yields were 7.2 nM and 7.8 nM using Illumina and KAPA methods (p>0.05), respectively. The average library yields from gDNA prepared from EDTA-blood of the same individuals were significantly greater using Illumina and KAPA methods (p<0.01). There was no difference between library yield with FTA or PC DBS with Illumina or KAPA methods (p>0.05; Table S2).

### Quality of whole genome sequences derived from DBS

The 63 genomic DNA libraries were sequenced on NovaSeq instruments with S1, S2, or S4 flow cells. Quality metrics of WGS from DBS and EDTA-blood were similar (Table S3 - S5). The average proportion of Q30 nucleotides of DBS libraries on S1, S2, and S4 flow cells were 93.7%, 92.9% and 89.5%, respectively, compared with 94.1%, 92.9%, and 87.8%, respectively, for the corresponding EDTA-blood libraries (p>0.05; Table S3). The average nucleotide error rates of DBS libraries on S1, S2, and S4 flow cells were 0.20%, 0.18%, and 0.22%, respectively, compared with 0.17%, 0.18%, and 0.27%, respectively, for the corresponding EDTA-blood libraries (p>0.05; Table S3).

The uniformity of WGS coverage in DBS was assessed by examining GC bias (defined as relative difference in average coverage of GC-rich regions (62%) to that of regions with the modal human genome GC-content (38%)). The range of GC bias required to pass quality control for clinical libraries made from blood samples was -0.25 to 0.25. 92.5% (49/53) of DBS WGS prepared with the Illumina method and 87.2% (34/39) DBS WGS prepared with the KAPA method were within this range (Table S4, S6). The uniformity of sequence coverage of WGS was also assessed by the standard deviation of coverage normalized to the average coverage and the total length of all sequenced reference genome nucleotides (Table S4, S7). There were no significant differences seen between fresh blood, manufactured DBS, or archived DBS in these measures (p>0.05). The average standard deviation of coverage normalized to average coverage was 0.19 for WGS from EDTA-blood (range 0.17-0.20, Table S4), 0.23 for WGS from manufactured DBS (range 0.19 – 0.37, Table S4), and 0.23 in California Department of Public Health DBS (range 0.19-0.27, Table S7). The average mappable genome length was 2.67 GB for all groups.

DNA damage (cytosine deamination causing C > T transitions) was assessed using several metrics: Firstly, an indirect measure was SNV concordance between WGS of fresh blood and DBS from the same individuals (Cohort 1, Table S8). It was over 92.6% (Table 3, S8). Secondly, there were no statistically significant differences in transition to transversion (Ti/Tv) ratio between WGS from DBS archived by the California Department of Public Health DBS (average 2.03 ± 0.01 [standard deviation], Table S7), fresh blood (2.03 ± 0.01) or manufactured DBS with Illumina or KAPA methods (2.03 ± 0.01; Table S4, S7). Thirdly, the rate of cytosine deamination was measured by calculating C>T+G>A / T>C+A>G variant ratios (Table S4, S7). There were no significant differences between WGS from fresh blood, manufactured DBS, or California Department of Public Health archived DBS.

**Table 3.**
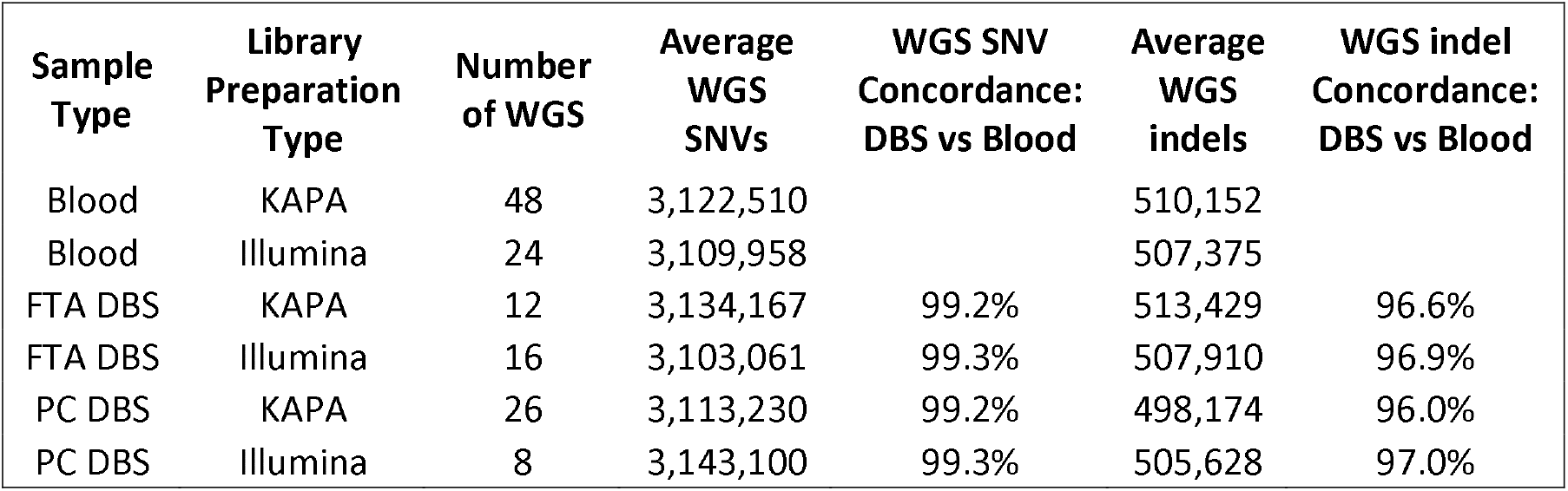
Concordance of SNVs and indels in WGS from 25 blood samples and 63 DBS derived from those samples. Libraries were prepared from two types of DBS cards (FTA and Protein Saver [PC]) using two preparation methods (Illumina PCR-free and KAPA HyperPlus).

### Quality of alignment and variant calling of whole genome sequences from DBS

Approximately 120 Gb of WGS was generated for 63 DBS-derived libraries and matched blood samples with the KAPA and Illumina library preparation methods. The average genomic coverage of WGS with KAPA was greater than that of Illumina (37.6-fold vs 43.2-fold, p<0.01; Table S4). However, the proportion of OMIM genes in which 100% of coding domain nucleotides had coverage ≥ 10X, an important measure of the ability to call heterozygous variants with confidence, did not differ between these methods (95.0% vs 93.6%, p>0.05; Table S4). The average genomic coverage of WGS for the corresponding EDTA-blood libraries was similar to that of the DBS-derived KAPA method (43.0-fold, p>0.05, Table S5). However, the proportion of OMIM genes in which 100% of coding domain nucleotides had ≥ 10X coverage was greater in the corresponding EDTA-blood libraries (96.6%, p<0.01; Table S5). There were not significant differences between FTA and PC DBS cards in either of these quality metrics using either library preparation method (p>0.05, Table S4, S5). As expected, the proportion of OMIM genes with ≥ 10X coverage of the complete coding domain increased with the depth of WGS (Figure 3). WGS with DBS-derived Illumina libraries and matched blood samples with the KAPA and Illumina methods had very similar distributions of proportions of OMIM genes with ≥ 10X coverage of the complete coding domain. However, DBS-derived KAPA libraries had much more variable distributions of proportions of OMIM genes with ≥ 10X coverage of the complete coding domain. Thus, DBS-derived KAPA libraries required generation of approximately 60 GB more WGS than DBS-derived Illumina libraries or blood-derived KAPA or Illumina libraries to provide ≥ 10X coverage of the complete coding domain of 92% of OMIM genes. SNV and nucleotide indel variant call accuracy was evaluated by comparing average SNV and indel concordance between WGS from DBS and blood in the same individuals. Mean concordance was 92.6% (range 87.2-94.6%) (Table S8). An average of 2.3% of variants were unique to blood samples, and 5.1% variants were unique to DBS (Table S8). We examined twenty random discordant calls in WGS from DBS and blood in a proband/sibling/father trio (Fig. S2-S16). Almost all discrepant variants occurred in regions that were either difficult to sequence, align, or variant call due to repetitive sequences (such as LINE1, Alu, FLAM, MER, or MStB1 endogenous retroviral elements), GC- or AT-rich or homopolymer-containing regions, or had more than one overlapping variant.

**Figure 3:**
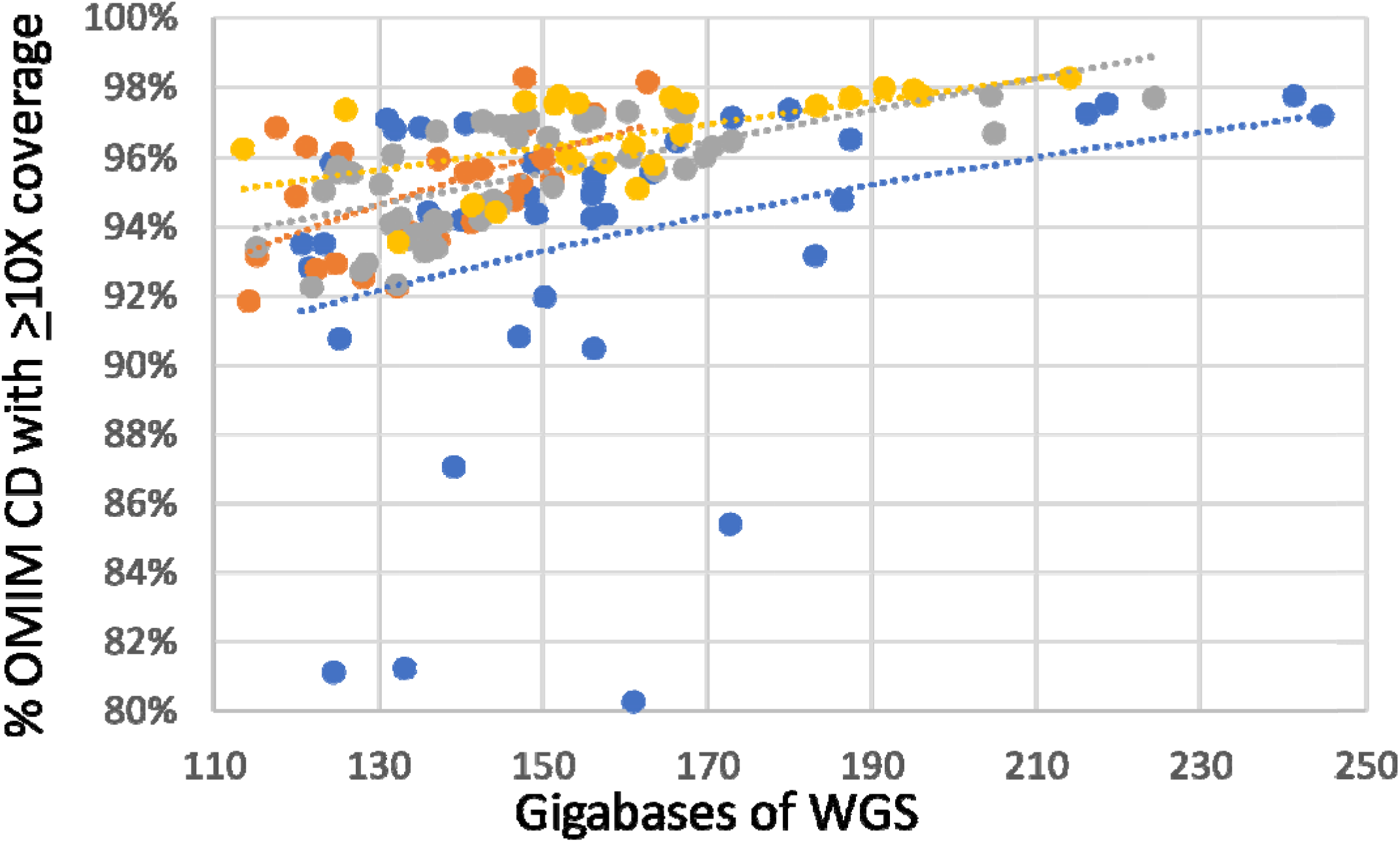
Relationship between percentage of MIM genes with at least 10-fold coverage of all coding domain nucleotides and amount of WGS. Shown are WGS from DBS prepared with the Illumina method (orange) or KAPA method (sky blue) and matched blood samples prepared with the Illumina method (grey) or KAPA method (yellow). Dotted trend lines are shown.

### Diagnostic performance of DBS-derived WGS

We evaluated the diagnostic recall of WGS performed on DBS in the 19 individuals who had at least one variant reported clinically (Table 1). These were assessed manually in the Integrated Genome Viewer (IGV), as well as by our standard annotation (Fabric genomics), variant calling and analysis pipeline (Table S1). All 13 SNVs, 2 indels, 9 SV-deletions and 4 SV-insertions were recapitulated in DBS-derived WGS (Table 1).

### WGS from DBS obtained from the California State Biobank

Having established that high quality WGS was possible from manufactured DBS, we sought to examine whether the same methods were effective with 29 randomly selected DBS samples obtained from the California State Biobank. These DBS were collected between 2/22/2000 – 04/06/2020. Six to twelve punches were available per sample (average 10, Table S6). Sixteen DBS were FTA, six were PerkinElmer filter paper, and nine DBS were of unknown type (Table S1). All DBS yielded high molecular weight DNA (mean 645 ng, range 331 – 1,050 ng, SD 189.74 ng, Table S6). The resultant gDNAs yielded sequencing libraries that passed quality control and produced high-quality WGS (Table S6). The average sequencing yield was 135 GB (SD 25) (Table S6). Resultant average genomic coverage was 38.6-fold (range 27.8 - 56.6, SD 7.0), while the average coverage of coding domains of OMIM genes was 36.4 (range 25.0 - 56.7, SD 7.0; Table S6). The average percentage of OMIM genes in which 100% of coding domain nucleotides had >10x coverage was 91.6% (Table S6). 59% (17/29) of the samples met our standard, clinical quality metric for this measure (>95% of OMIM genes with >10x coverage).

### Effect of DBS Age and Paper Type

Statistical analyses were performed on the second cohort of archived DBS WGS to investigate the potential effects of DBS sample age and paper type used. There was no significant correlation between age of the DBS sample and WGS yield, genome coverage, coverage of OMIM genes, or mitochondrial coverage (Figure S1). Nor was there a significant effect of filter paper type on WGS yield or genome coverage. In a previous study, the amount of input DNA had affected the proportion of duplicate reads and the overall coverage of WES.^20^ We therefore investigated the potential correlation using Pearson’s product-moment correlation. We found no significant correlation between amount of input DNA and proportion of duplicate reads or overall coverage (Table S2, S6).

## DISCUSSION

Archived DBS from national and state screening programs worldwide present an enormous resource for investigating the etiology of pediatric genetic diseases. Here, we demonstrated that high-quality WGS data can be obtained from archived DBS stored for up to 20 years. We also demonstrated in a panel of samples that WGS from DBS identified all of the SNV, indel, SV-deletion and SV-duplication findings that had been reported clinically from prior WGS from blood samples, as well as a 92.6% concordance of SNV and small indel calls between WGS from blood and DBS.

The methods described herein are simple and use generally available kits (Supplementary Methods). They will enable researchers and clinical laboratories to utilize DBS for PCR-free WGS. DNA isolation from DBS took less than 90 minutes. Very little degradation of DNA extracted from DBS was observed by agarose gel electrophoresis (Figure 1). DNA purity (A260/A280 ratio) was slightly lower than that from whole blood but acceptable for WGS, as confirmed by secondary WGS QC metrics and tertiary analysis results.

Avoidance of PCR is important for optimal analytic performance of WGS, particular for SVs^28–30^. Compared with gDNA derived from EDTA blood from the same individuals, the library yields of DBS were lower but the primary, secondary, and tertiary analysis of WGS passed quality control criteria. The average coverage of OMIM genes was 36.0x and 42.6x for the Illumina and KAPA preparation methods, respectively, in samples analyzed at RCIGM, and 36.4x in DBS obtained from the California Biobank. This coverage supports confident heterozygous variant calling. There was no observed difference in the quality of WGS associated with sample age or DBS filter paper type. While both the KAPA and Illumina methods worked well, there was much greater variability in the proportion of OMIM genes with at least 10-fold coverage of their coding domain nucleotides with the former. Thus the Illumina method more consistently yielded adequate disease gene coverage, which is important in clinical production WGS.

The ability to perform high quality PCR-free WGS from archived DBS is significant for several reasons. First, there are over 18 million samples in the California Biobank that encompass a full range of ethnic, socioeconomic, regional, and temporal diversity within the state. This represents an enormous resource for genomic studies. Large-scale sequencing studies of these DBS have the potential to reveal new genotype-phenotype associations and further our knowledge of genetic diseases in human populations. Currently, consent for sample storage is obtained at the time of collection, and these DBS samples are then stored indefinitely in the California State Biobank. This wealth of stored biospecimen data may support large-scale studies to investigate the epidemiology of rare genetic disease in a way that has not previously been feasible. For example, the San Diego Study of Outcomes for Mothers and Infants (SOMI) dataset currently supports linkage of vital statistics, DBS samples, death certificates and hospital records for infants born in San Diego County after 2007. These types of datasets will enable the linkage of epidemiological data with individual-level molecular diagnoses.

Second, there is growing interest in the potential of WGS for expanded newborn screening.^12,20,31^ Advances in WGS technology have recently made fully automated, diagnostic WGS possible in 19.5 hours.^21^ Those methods are compatible with the WGS from DBS described herein. Thus, autonomous, expanded newborn screening by WGS of DBS is conceivable. In screening mode, WGS interpretation would be limited to known pathogenic and likely pathogenic variants, which could be detected without the need for trio sequencing - confirmatory testing could incorporate parental samples if warranted, as is done currently. Adding new conditions to the recommended uniform screening panel is costly, and each additional assay requires independent state or federal legislative approval.^32^ In California, 84 conditions are currently screened and 1 in 600 newborns has a positive NBS result.^11^ In contrast, WGS has the potential to screen for known pathogenic and likely pathogenic variants in over 6,000 disorders at once, including those that can cause sudden infant death.^14,15,33^ There are many rare genetic diseases, such as pyridoxine-dependent epilepsy, which may have effective treatments and meet the Wilson and Junger criteria for inclusion in newborn screening.^34,35^ Newborn screening by WGS has potential to reduce morbidity and mortality associated with these conditions. Certain genetic diseases, such as spinal muscular atrophy, also rely on the particular variants identified to guide specific management. As shown herein in two cases, WGS provides this level of analytic performance. Here, we have demonstrated that PCR-free WGS is feasible for use on archived DBS collected from NBS, expanding the potential of their use for investigating the prevalence of rare genetic diseases.

## MATERIALS AND METHODS

### Study Design

This study received a waiver of consent from the Rady Children’s Hospital and University of California – San Diego (UCSD) institutional review boards (IRB) and was undertaken as a quality improvement project. Samples were from two cohorts: The first consisted of 25 individuals who received WGS from whole blood at the Rady Children’s Institute for Genomic Medicine (RCIGM) between January 2018 and June 2019 for diagnosis of a suspected genetic disease. WGS was performed in laboratories accredited by the College of American Pathologists (CAP) and certified through Clinical Laboratory Improvement Amendments (CLIA). They had either been consented under various research protocols approved by the UCSD IRB and the Johns Hopkins IRB or were sample retains from clinical diagnostic testing. Blood spots were created using two paper types (filter chemical treated and non-treated) and stored for these individuals. Two cases had additional family members available for analysis: sample 6 had an affected sibling and father available, and sample 8 had father’s genome available (Table 1). WGS data from DBS were compared with prior, clinical WGS data from whole blood. The second cohort consisted of 29 randomly selected, de-identified, anonymized DBS obtained from the California Biobank Program. Two spots were collected for each sample, and a variable number of punches was available (Table S6). All samples had >7 punches available, and 6 punches were actually used for each sample to maintain consistency (Table S1 and S6). Samples were archived after collection and stored with desiccant at -20^°^C. They represented a range of filter types and years in storage. No IRB approval was required from the CDPH IRB for the use of randomly selected DBS samples with no associated clinical data to determine feasibility of methods.

### Preparation of DBS using EDTA whole blood

Twenty-nine DBS sets (each set containing multiple cycles or spots, see Table S1 for details) were prepared with Whatman NUCLEIC-CARD™ matrix (FTA) (ThermoFisher, Catalog #: 4473975) and 34 DBS sets were prepared with 903 Protein Saver 903 Cards (PC) (GE Healthcare, Catalog #:10534612) from the 25 individuals. Each DBS was made with 40 µl EDTA-whole blood. Multiple spots were made per individual (Table S1).

### Genomic DNA isolation and sequencing library preparation

DBS were lysed according to either of two lysis protocols using either six or ten punches from DBS (Supplementary Methods). Additionally, a titration experiment was performed with inputs of 1, 3, 5, 7, or 10 DBS punches (3mm^2^) per extraction (Supplementary materials). Genomic DNA was isolated with DNA Flex Lysis Reagent Kit (Illumina) or Proteinase K (QIAGEN). The gDNA quality was assessed with the Quant-iT Picogreen dsDNA Assay Kit (ThermoFisher) and Nanodrop A260/A280 assays (ThermoFisher).^36,37^ The integrity of extracted gDNA was examined by electrophoresis using 0.8% agarose gels (ThermoFisher). PCR-free libraries were prepared with either DNA PCR-free (Tagmentation) Prep kits (Illumina) or KAPA HyperPlus PCR-free library kits (Roche, abbreviated KAPA herein) for the 63 manufactured DBS, according to the manufacturer’s instructions (Table S1).^38,39^ For the archived DBS from California Department of Public Health (CDPH), WGS libraries were prepared using Illumina prep kits.

### Whole genome sequencing

Libraries with concentration >3 nM and acceptable fragment size (KAPA Hyper kit) passed quality control and were sequenced on Illumina Novaseq 6000 instruments. Libraries from the 63 manufactured DBS were pooled at equal molarity as follows: S1 flow cell, 2.5 libraries, S2 flow cell, 5 – 6 libraries, or S4 flow cell, 24 libraries. Quality metrics for passing a WGS were Q30≥ 80%, error rate ≤ 3% and >120 Gb per WGS (see Supplementary Methods).

### Secondary analysis of whole genome sequences

WGS from the manufactured DBS were aligned to human genome assembly GRCh37 (hg19) and variants identified with the Illumina DRAGEN (Dynamic Read Analysis for GENomics) Bio-IT Platform (Illumina, Table S1). WGS from archived DBS were aligned to human genome assembly GrCh37 (hg19) and nucleotide variants were identified with the DRAGEN platform (Illumina, Table S1). Samples were run using different versions of DRAGEN as described in Table S1. For each update of DRAGEN, a verification process was applied to ensure the quality of DRAGEN variant calling as it pertains to the RCIGM clinical diagnostic standards. Briefly, for each DRAGEN upgrade, VCF concordance between old and new DRAGEN results are verified to have >99% F2-measure as calculated by vcfeval.^40^ Furthermore, quality control metrics, such as mapping rate and average genomic coverage are verified to have >98% concordance between old and new DRAGEN results. Structural variants were identified with Manta and CNVnator (using DNAnexus). Structural variants were filtered to retain those affecting coding regions of known disease genes and with allele frequencies <2% in the RCIGM database. All samples underwent a battery of quality controls, including: 1) sample identity tracking (STR/CODIS) from orthogonal inputs (capillary electrophoresis using Genetic Analyzer ThermoFisher 3500xl) and in silico STR from WGS; 2) <15% duplicate rate, 3) >98% aligned reads rate; 4) Ti/Tv in appropriate range (2.0-2.2); 5) Hom/Het in appropriate range (0.50-0.61); 6) >90% of OMIM genes with >10-fold coverage of every coding nucleotide; 7) sex match; 8) additional technical controls (insert size and others). Coverage uniformity was assessed using the GC bias measure as well as two additional measures: the standard deviation of coverage normalized to average coverage and the total length of all reference genome regions with read coverage. Both of these measures were computed by binning the complete genome coverage into bins of 200 bases at a time (Table S4 and S7). For a small fraction of the sample, these metrics were not retained, and these have been marked as ND (Table S4 and S7).

### Surveillance of cross-sample contamination during DBS WGS process

Intra-sample and intra-batch cross contamination were monitored with following measurements: 1. Ensure cleanness of each DBS sample when using a Harris Uni-Core punch to remove a sample disc to a sample tube by punching on a clean paper before working on the next DBS sample according to manufacturer’s recommendation. 2. A negative control (NTC) was included during WGS library construction and QC quantification, samples in a batch with NTC contamination would be failed and would not be passed for sequencing. 3. In silico analysis post sequencing, cross-sample contamination was computed and had to pass defined criteria for downstream variant calling.

### Concordance Analysis between EDTA blood and DBS WGS

Small nucleotide variants concordance analysis for datasets derived from EDTA blood samples and DBS samples were performed according to best practices set forth by Global Alliance for Genomics and Health (GA4GH) Benchmarking Team.^41^ Briefly, after generation, the VCF files were compared using the vcfeval software.^41^ The GA4GH Benchmarking Team developed standardized performance metrics for genomic variant calls as well as sophisticated variant comparison tools to robustly compare different representations of the same variant, and a set of standard browser extensible data (BED) files describing difficult genome contexts to stratify performance. The GA4GH Benchmarking application requires a truth VCF file (for this study, the EDTA blood sample), the truth confident regions (the GIAB high-confidence BED file for HG002 was used), and the query VCF file (the DBS sample in this study). The GA4GH application returns the count of false negatives (FNs), false positives (FPs), and true positives (TPs) in both standardized VCF and comma-separated value formats. Performance metrics follow the GA4GH standardized definitions, in which genotyping errors are counted both as FP and FN. Precision (also known as positive prediction value (PPV)) was calculated using the following formula: PPV = TP/(TP + FP). Sensitivity was calculated using the following formula: sensitivity = TP/(TP + FN).

### Diagnostic utility of WGS from DBS

Nineteen of the 25 individuals in whom DBS were manufactured had received diagnostic results from clinical WGS from whole blood at RCIGM according to American College of Medical Genetics and Genomics (ACMG)/Association of Molecular Pathology (AMP) guidelines. Aligned sequences from DBS-based WGS were viewed in the Integrated Genomics Viewer and Fabric Genomics to determine whether the variants and diplotypes were recapitulated.

### Statistical Analysis

A student’s t-test was used to compare the means of test and control groups. P values <0.05 were considered statistically significant. Pearson product moment correlations were used to analyze relationships between age of the bloodspot and quality metrics. Two-tailed Fisher’s exact tests were performed to compare the effects of different paper types. All analyses were conducted in R v.4.0.3, and visualization was done using the packages ggpubr and ggplot2.^42,43^

## Data Availability

All data produced in the present study are available upon reasonable request to the authors

## ACKNOWLEDGMENTS

This work was supported by grant HD101540 from NICHD.

## DATA DEPOSITION AND ACCESS

These data are subject to conditions of the IRB protocols and CDPH policies under which the data was generated, and therefore the raw sequencing data is unavailable. For additional summary or aggregate level data, please contact Dr. Stephen Kingsmore.

## AUTHOR CONTRIBUTIONS

YD, MO, SN and SK designed the study. YD, SB, ZBO, LVDK, JL and ZZ performed sequencing and analysis. YD, SK and MO wrote the manuscript and made the figures and tables. NV, SB, KH and KC performed bioinformatic analyses. SB and MB aided in study design and data analysis. RB, GB and CC obtained DBS from the California State Biobank and provided logistical and analytic support. All authors reviewed the final version. SK and CC acted as the principal investigator on the study, edited the manuscript, and contributed to study design along with JG.

