## Supplementary Materials for "Scalable, high quality, whole genome sequencing from archived, newborn, dried blood spots"

**Supplementary Material**

**Supplementary Tables**

**Table S1. Sample characteristics and number of DBS made for each sample.** Abbreviations: FTA: FTA card; PC: protein saver card; KAPA: KAPA Hyper Plus library method; Qiagen: Qiagen lysis method for DNA isolation; Illumina: Tagmentation PCR-free library method; #: the spot from CDPH was double size of simulated DBS.

| **Sample ID** | **Simulated DBS or CDPH DBS?** | **Number of simulated DBS made** | **Number of DBS punches used for Isolation** | **Age of DBS at gDNA isolation** | **DBS paper type** | **DNA extraction method** | **Library preparation method** | **Quantity of DNA used for library preparation (ng)** | **DRAGEN version** |
| --- | --- | --- | --- | --- | --- | --- | --- | --- | --- |
| Subject 1 | Simulated DBS | 8 | 6 | 52 | FTA | Illumina | Illumina | 267.60 | 2.1.5 |
| Subject 1 | Simulated DBS |  | 10 | 58 | FTA | Qiagen | KHP | 420.90 | 2.1.5 |
| Subject 1 | Simulated DBS | 10 | 6 | 52 | PC | Illumina | Illumina | 246.60 | 2.1.5 |
| Subject 1 | Simulated DBS |  | 10 | 8 | PC | Qiagen | KHP | 300.00 | 2.1.5 |
| Subject 1 | Simulated DBS |  | 10 | 37 | PC | Qiagen | KHP | 526.65 | 2.1.5 |
| Subject 10 | Simulated DBS | 2 | 6 | 52 | FTA | Illumina | Illumina | 169.20 | 2.1.5 |
| Subject 10 | Simulated DBS |  | 10 | 58 | FTA | Qiagen | KHP | 481.50 | 2.1.5 |
| Subject 10 | Simulated DBS | 3 | 6 | 52 | PC | Illumina | Illumina | 210.60 | 2.1.5 |
| Subject 10 | Simulated DBS |  | 10 | 8 | PC | Qiagen | KHP | 300.00 | 2.1.5 |
| Subject 10 | Simulated DBS |  | 10 | 58 | PC | Qiagen | KHP | 339.18 | 2.1.5 |
| Subject 11 | Simulated DBS | 8 | 6 | 52 | FTA | Illumina | Illumina | 500.00 | 2.1.5 |
| Subject 11 | Simulated DBS |  | 6 | 64 | FTA | Illumina | Illumina | 283.20 | 2.1.5 |
| Subject 11 | Simulated DBS |  | 10 | 183 | FTA | Qiagen | KHP | 522.80 | 2.1.5 |
| Subject 11 | Simulated DBS | 10 | 6 | 53 | PC | Illumina | Illumina | 500.00 | 2.1.5 |
| Subject 11 | Simulated DBS |  | 10 | 9 | PC | Qiagen | KHP | 300.00 | 2.1.5 |
| Subject 11 | Simulated DBS |  | 10 | 38 | PC | Qiagen | KHP | 380.98 | 2.1.5 |
| Subject 12 | Simulated DBS | 4 | 10 | 37 | FTA | Qiagen | KHP | 571.17 | 2.1.5 |
| Subject 12 | Simulated DBS | 2 | 10 | 8 | PC | Qiagen | KHP | 300.00 | 2.1.5 |
| Subject 12 | Simulated DBS |  | 10 | 37 | PC | Qiagen | KHP | 357.46 | 2.1.5 |
| Subject 13 | Simulated DBS | 4 | 6 | 52 | FTA | Illumina | Illumina | 500.00 | 2.1.5 |
| Subject 13 | Simulated DBS |  | 6 | 64 | FTA | Illumina | Illumina | 501.30 | 2.1.5 |
| Subject 13 | Simulated DBS | 2 | 6 | 37 | PC | Illumina | Illumina | 500.00 | 2.1.5 |
| Subject 13 | Simulated DBS |  | 10 | 9 | PC | Qiagen | KHP | 300.00 | 2.1.5 |
| Subject 14 | Simulated DBS | 4 | 6 | 52 | FTA | Illumina | Illumina | 206.70 | 2.1.5 |
| Subject 14 | Simulated DBS | 2 | 6 | 52 | PC | Illumina | Illumina | 186.90 | 2.1.5 |
| Subject 14 | Simulated DBS |  | 10 | 8 | PC | Qiagen | KHP | 300.00 | 2.1.5 |
| Subject 15 | Simulated DBS | 2 | 10 | 74 | FTA | Qiagen | KHP | 300.00 | 2.1.5 |
| Subject 15 | Simulated DBS | 2 | 10 | 74 | PC | Qiagen | KHP | 300.00 | 2.1.5 |
| Subject 16 | Simulated DBS | 2 | 10 | 99 | PC | Qiagen | KHP | 276.63 | 2.1.5 |
| Subject 17 | Simulated DBS | 2 | 10 | 77 | PC | Qiagen | KHP | 279.75 | 2.1.5 |
| Subject 18 | Simulated DBS | 2 | 6 | 6 | FTA | Illumina | Illumina | 245.70 | 2.1.5 |
| Subject 18 | Simulated DBS | 2 | 6 | 6 | PC | Illumina | Illumina | 301.80 | 2.1.5 |
| Subject 19 | Simulated DBS | 2 | 6 | 6 | FTA | Illumina | Illumina | 171.90 | 2.1.5 |
| Subject 19 | Simulated DBS | 2 | 6 | 6 | PC | Illumina | Illumina | 177.60 | 2.1.5 |
| Subject 2 | Simulated DBS | 2 | 10 | 58 | FTA | Qiagen | KHP | 1003.80 | 2.1.5 |
| Subject 2 | Simulated DBS | 2 | 10 | 8 | PC | Qiagen | KHP | 300.00 | 2.1.5 |
| Subject 2 | Simulated DBS |  | 10 | 58 | PC | Qiagen | KHP | 852.90 | 2.1.5 |
| Subject 20 | Simulated DBS | 2 | 6 | 439 | FTA | Illumina | Illumina | 178.50 | 2.1.5 |
| Subject 21 | Simulated DBS | 2 | 6 | 397 | FTA | Illumina | Illumina | 228.00 | 2.1.5 |
| Subject 22 | Simulated DBS | 2 | 6 | 380 | FTA | Illumina | Illumina | 214.50 | 2.1.5 |
| Subject 23 | Simulated DBS | 2 | 6 | 636 | FTA | Illumina | Illumina | 154.80 | 2.1.5 |
| Subject 24 | Simulated DBS | 2 | 6 | 661 | FTA | Illumina | Illumina | 191.10 | 2.1.5 |
| Subject 25 | Simulated DBS | 2 | 6 | 122 | FTA | Illumina | Illumina | 199.20 | 2.1.5 |
| Subject 3 | Simulated DBS | 2 | 10 | 122 | FTA | Qiagen | KHP | 738.30 | 2.1.5 |
| Subject 3 | Simulated DBS | 2 | 10 | 87 | PC | Qiagen | KHP | 300.00 | 2.1.5 |
| Subject 3 | Simulated DBS |  | 10 | 101 | PC | Qiagen | KHP | 369.62 | 2.1.5 |
| Subject 4 | Simulated DBS | 4 | 6 | 104 | FTA | Illumina | Illumina | 312.00 | 2.1.5 |
| Subject 4 | Simulated DBS |  | 10 | 110 | FTA | Qiagen | KHP | 587.40 | 2.1.5 |
| Subject 4 | Simulated DBS | 2 | 6 | 104 | PC | Illumina | Illumina | 417.00 | 2.1.5 |
| Subject 4 | Simulated DBS |  | 10 | 75 | PC | Qiagen | KHP | 300.00 | 2.1.5 |
| Subject 5 | Simulated DBS | 2 | 10 | 110 | FTA | Qiagen | KHP | 675.90 | 2.1.5 |
| Subject 5 | Simulated DBS | 2 | 10 | 110 | PC | Qiagen | KHP | 308.38 | 2.1.5 |
| Subject 5 | Simulated DBS |  | 10 | 75 | PC | Qiagen | KHP | 300.00 | 2.1.5 |
| Subject 6 | Simulated DBS | 8 | 10 | 102 | FTA | Qiagen | KHP | 300.00 | 3.4.5 |
| Subject 6 | Simulated DBS | 10 | 10 | 75 | PC | Qiagen | KHP | 300.00 | 3.4.5 |
| Subject 6 | Simulated DBS |  | 10 | 89 | PC | Qiagen | KHP | 312.84 | 3.4.5 |
| Subject 7 | Simulated DBS | 2 | 10 | 60 | FTA | Qiagen | KHP | 565.20 | 2.1.5 |
| Subject 7 | Simulated DBS | 2 | 10 | 25 | PC | Qiagen | KHP | 300.00 | 2.1.5 |
| Subject 7 | Simulated DBS |  | 10 | 10 | PC | Qiagen | KHP | 300.00 | 2.1.5 |
| Subject 8 | Simulated DBS | 8 | 10 | 39 | FTA | Qiagen | KHP | 300.00 | 2.1.5 |
| Subject 8 | Simulated DBS | 10 | 10 | 18 | PC | Qiagen | KHP | 220.28 | 2.1.5 |
| Subject 9 | Simulated DBS | 8 | 10 | 36 | FTA | Qiagen | KHP | 519.00 | 2.1.5 |
| Subject 9 | Simulated DBS | 10 | 10 | 1 | PC | Qiagen | KHP | 300.00 | 2.1.6 |
| 1 | CDPH DBS | 1 | 6 | 167 | Whatman | Illumina | Illumina | 425.25 | 3.5.7 |
| 2 | CDPH DBS | 1 | 6 | 167 | Whatman | Illumina | Illumina | 500.00 | 3.5.7 |
| 3 | CDPH DBS | 1 | 6 | 180 | Whatman | Illumina | Illumina | 316.23 | 3.5.7 |
| 4 | CDPH DBS | 1 | 6 | 193 | Whatman | Illumina | Illumina | 485.00 | 3.5.7 |
| 5 | CDPH DBS | 1 | 6 | 524 | Whatman | Illumina | Illumina | 486.25 | 3.5.7 |
| 6 | CDPH DBS | 1 | 6 | 675 | Whatman | Illumina | Illumina | 498.65 | 3.5.7 |
| 7 | CDPH DBS | 1 | 6 | 2011 | Whatman | Illumina | Illumina | 258.00 | 3.5.7 |
| 8 | CDPH DBS | 1 | 6 | 2011 | Whatman | Illumina | Illumina | 267.75 | 3.5.7 |
| 9 | CDPH DBS | 1 | 6 | 2250 | Whatman | Illumina | Illumina | 358.80 | 3.5.7 |
| 10 | CDPH DBS | 1 | 6 | 2267 | Whatman | Illumina | Illumina | 460.85 | 3.5.7 |
| 11 | CDPH DBS | 1 | 6 | 2518 | PerkinElmer | Illumina | Illumina | 544.88 | 3.5.7 |
| 12 | CDPH DBS | 1 | 6 | 2536 | PerkinElmer | Illumina | Illumina | 500.00 | 3.5.7 |
| 13 | CDPH DBS | 1 | 6 | 2742 | PerkinElmer | Illumina | Illumina | 429.65 | 3.5.7 |
| 14 | CDPH DBS | 1 | 6 | 2742 | PerkinElmer | Illumina | Illumina | 500.00 | 3.5.7 |
| 15 | CDPH DBS | 1 | 6 | 3297 | Whatman | Illumina | Illumina | 473.78 | 3.5.7 |
| 16 | CDPH DBS | 1 | 6 | 3297 | Whatman | Illumina | Illumina | 454.75 | 3.5.7 |
| 17 | CDPH DBS | 1 | 6 | 4661 | Whatman | Illumina | Illumina | 361.38 | 3.5.7 |
| 18 | CDPH DBS | 1 | 6 | 4661 | Whatman | Illumina | Illumina | 500.00 | 3.5.7 |
| 19 | CDPH DBS | 1 | 6 | 4720 | Whatman | Illumina | Illumina | 433.73 | 3.5.7 |
| 20 | CDPH DBS | 1 | 6 | 4720 | Whatman | Illumina | Illumina | 377.70 | 3.5.7 |
| 21 | CDPH DBS | 1 | 6 | 5862 | Unknown | Illumina | Illumina | 268.00 | 3.5.7 |
| 22 | CDPH DBS | 1 | 6 | 5988 | Unknown | Illumina | Illumina | 475.20 | 3.5.7 |
| 23 | CDPH DBS | 1 | 6 | 5971 | Unknown | Illumina | Illumina | 346.60 | 3.5.7 |
| 24 | CDPH DBS | 1 | 6 | 6429 | Unknown | Illumina | Illumina | 500.00 | 3.5.7 |
| 25 | CDPH DBS | 1 | 6 | 6429 | Unknown | Illumina | Illumina | 500.00 | 3.5.7 |
| 26 | CDPH DBS | 1 | 6 | 6524 | Unknown | Illumina | Illumina | 500.00 | 3.5.7 |
| 27 | CDPH DBS | 1 | 6 | 6524 | Unknown | Illumina | Illumina | 500.00 | 3.5.7 |
| 28 | CDPH DBS | 1 | 6 | 7516 | Unknown | Illumina | Illumina | 500.00 | 3.5.7 |
| 29 | CDPH DBS | 1 | 6 | 7516 | Unknown | Illumina | Illumina | 500.00 | 3.5.7 |

**Table S2: Quantity and quality of genomic DNA and sequencing library concentration of 63 manufactured DBS from 25 individuals, utilizing two types of filter papers, compared with matched blood samples.** NA: not applicable.

| **Subject** |  | **FTA Dried Blood Spot** | | | | **Protein Saver Dried Blood Spot** | | | | **350 µl Blood Sample** | | | |
| --- | --- | --- | --- | --- | --- | --- | --- | --- | --- | --- | --- | --- | --- |
|  | **Library Preparation Method** | **DNA yield (ng)** | **A260/ A280 ratio** | **Library input DNA (ng)** | **Library yield (nM)** | **DNA yield (ng)** | **A260/ A280 ratio** | **Library input DNA (ng)** | **Library yield (nM)** | **DNA yield (ng)*** | **A260/ A280 ratio** | **Library input DNA (ng)** | **Library yield (nM)** |
| 1 | Illumina | 285 | 1.59 | 268 | 7.7 | 263 | 1.54 | 247 | 8.8 | 6,670 | 1.85 | 1000 | 13.4 |
| 4 |  | 333 | 1.61 | 312 | 7.3 | 445 | 1.52 | 417 | 7.1 | 7,340 | 1.83 | 1000 | 14.4 |
| 10 |  | 180 | 1.48 | 169 | 5.9 | 225 | 1.44 | 211 | 6.9 | 5,332 | 1.78 | 1000 | 13.9 |
| 11_1 |  | 844 | 1.64 | 500 | 10.3 | 920 | 1.67 | 500 | 10.2 | 9,244 | 1.82 | 1000 | 19.2 |
| 13_1 |  | 761 | 1.77 | 500 | 8.4 | 674 | 1.82 | 500 | 8.1 | 13,130 | 1.84 | 1000 | 14.4 |
| 14 |  | 220 | 1.48 | 207 | 5.7 | 199 | 1.46 | 187 | 7.6 | 5,580 | 1.8 | 1000 | 27.3 |
| 18 |  | 262 | 1.42 | 246 | 5.9 | 322 | 1.46 | 302 | 7.1 | 5,096 | 1.8 | 1000 | 10.7 |
| 19 |  | 183 | 1.48 | 172 | 4.9 | 189 | 1.53 | 178 | 4.6 | 4,672 | 1.8 | 1000 | 11.8 |
| 20 |  | 190 | 1.6 | 179 | 6.6 | NA | NA | NA | NA | 7,200 | 1.71 | 1000 | 13.2 |
| 21 |  | 243 | 1.59 | 228 | 7.8 | NA | NA | NA | NA | 9,537 | 1.79 | 1000 | 20.3 |
| 22 |  | 229 | 1.7 | 215 | 6.6 | NA | NA | NA | NA | 6,046 | 1.65 | 1000 | 13.4 |
| 23 |  | 165 | 1.54 | 155 | 7 | NA | NA | NA | NA | 6,831 | 1.81 | 1000 | 15.3 |
| 24 |  | 204 | 1.53 | 191 | 6.6 | NA | NA | NA | NA | 9,521 | 1.82 | 1000 | 11.3 |
| 25 |  | 212 | 1.58 | 199 | 6.8 | NA | NA | NA | NA | 7,551 | 1.78 | 1000 | 15 |
| 11_2 |  | 302 | 1.59 | 283 | 5.9 | NA | NA | NA | NA | 9,244 | 1.82 | 1000 | 19.2 |
| 13_2 |  | 535 | 1.73 | 501 | 6.7 | NA | NA | NA | NA | 13,130 | 1.84 | 1000 | 14.4 |
| **Average** |  | **322** | **1.58** | **270** | **6.9** | **405** | **1.56** | **318** | **7.6** | **7,411** | **1.79** | **1000** | **15.3** |
| 1 | KAPA | 561 | 1.6 | 421 | 7.8 | 609 | 1.68 | 300 | 3 | 6,670 | 1.85 | 1000 | 13.4 |
| 2 |  | 1,338 | 1.75 | 1004 | 13.6 | 948 | 1.76 | 300 | 4.4 | 5,900 | 1.84 | 1000 | 12 |
| 3 |  | 984 | 1.76 | 738 | 9.2 | 1,728 | 1.72 | 300 | 4.8 | 10,240 | 1.84 | 1000 | 17.6 |
| 4 |  | 783 | 1.77 | 587 | 8.5 | 1,120 | 1.72 | 300 | 13.8 | 7,340 | 1.83 | 1000 | 14.4 |
| 5 |  | 901 | 1.77 | 676 | 5.6 | 987 | 1.72 | 308 | 3.7 | 5,190 | 1.79 | 1000 | 15.9 |
| 6 |  | 851 | 1.78 | 300 | 5.4 | 832 | 1.78 | 300 | 4.9 | 5,090 | 1.84 | 1000 | 15 |
| 7 |  | 754 | 1.75 | 565 | 6.1 | 694 | 1.76 | 300 | 6.4 | 5,260 | 1.85 | 1000 | 13.2 |
| 8 |  | 400 | 1.75 | 300 | 5.8 | 441 | 1.67 | 220 | 7.5 | 4,767 | 1.73 | 1000 | 13.8 |
| 9 |  | 692 | 1.63 | 519 | 7.3 | 792 | 1.53 | 300 | 6.6 | 6,115 | 1.78 | 1000 | 11.3 |
| 10 |  | 642 | 1.61 | 482 | 11.8 | 430 | 1.6 | 300 | 3.1 | 5,332 | 1.78 | 1000 | 13.9 |
| 11 |  | 1,394 | 1.72 | 523 | 18.8 | 1,638 | 1.76 | 300 | 7.1 | 9,244 | 1.82 | 1000 | 19.2 |
| 12 |  | 762 | 1.71 | 571 | 11.6 | 756 | 1.76 | 300 | 6.1 | 7,851 | 1.99 | 1000 | 23.9 |
| 13 |  | NA | NA | NA | NA | 1,312 | 1.82 | 300 | 15.9 | 13,130 | 1.84 | 1000 | 14.4 |
| 14 |  | NA | NA | NA | NA | 417 | 1.61 | 300 | 7.1 | 5,580 | 1.8 | 1000 | 27.3 |
| 15 |  | 409 | 1.66 | 300 | 7.2 | 624 | 1.66 | 300 | 7.1 | 5,356 | 1.83 | 1000 | 13.3 |
| 16 |  | NA | NA | NA | NA | 369 | 1.6 | 277 | 7.3 | 6,200 | 1.81 | 1000 | 11.8 |
| 17 |  | NA | NA | NA | NA | 373 | 1.77 | 280 | 12.2 | 6,370 | 1.8 | 1000 | 12.8 |
| **Average** |  | **806** | **1.71** | **537** | **9.1** | **828** | **1.7** | **293** | **7.1** | **6,583** | **1.83** | **1000** | **15.3** |
| 1 | KAPA |  |  |  |  | 702 | 1.83 | 527 | 7.4 | 6,670 | 1.85 | 1000 | 13.4 |
| 2 |  |  |  |  |  | 1,137 | 1.88 | 853 | 6.8 | 5,900 | 1.84 | 1000 | 12 |
| 3 |  |  |  |  |  | 1,971 | 1.8 | 370 | 5.1 | 10,240 | 1.84 | 1000 | 17.6 |
| 5 |  |  |  |  |  | 912 | 1.73 | 300 | 10.5 | 5,190 | 1.79 | 1000 | 15.9 |
| 6 |  |  |  |  |  | 1,251 | 1.79 | 313 | 6.2 | 5,090 | 1.84 | 1000 | 15 |
| 7 |  |  |  |  |  | 651 | 1.8 | 300 | 7.1 | 5,260 | 1.85 | 1000 | 13.2 |
| 10 |  |  |  |  |  | 452 | 1.65 | 339 | 13.2 | 5,332 | 1.78 | 1000 | 13.9 |
| 11 |  |  |  |  |  | 1,524 | 1.78 | 381 | 4.9 | 9,244 | 1.82 | 1000 | 19.2 |
| 12 |  |  |  |  |  | 715 | 1.72 | 357 | 4.5 | 7,851 | 1.99 | 1000 | 23.9 |
| **Average** |  |  |  |  |  | **1,035** | **1.78** | **416** | **7.3** | **6,753** | **1.84** | **1000** | **16** |

**Table S3:** **Quantity and quality of WGS derived from manufactured DBS and matched blood samples with two different library preparation methods (Illumina and KAPA) and three different sequencing flow cells.** Abbreviations: Q: quality score.

|  | **Library Preparation Method** | **Flowcell type** | **Number of flowcells used** | **Number of Samples for which WGS performed** | **Read Length (nt)** | **Total yield per flowcell (Gb)** | **Error rate (%)** | **% of Clusters passing filters** | **% of called bases with >Q30** | **% with Correct Index** |
| --- | --- | --- | --- | --- | --- | --- | --- | --- | --- | --- |
| **DBS research WGS** | **KAPA** | S1 | 15 | 36 | 2x101 | 415 | 0.2 | 75.3 | 93.7 | 94.0 |
|  |  | S2 | 10 | 49 | 2x101 | 947 | 0.2 | 76.8 | 92.9 | 93.8 |
|  | **Illumina** | S4 | 1 | 24 | 2x101 | 3270 | 0.2 | 67.2 | 89.5 | 86.0 |
| **Clinical grade WGS (blood samples)** | **KAPA** | S1 | 8 | 20 | 2x101 | 444 | 0.2 | 80.8 | 94.1 | 95.3 |
|  |  | S2 | 5 | 28 | 2x101 | 977 | 0.2 | 78.7 | 92.9 | 95.1 |
|  | **Illumina** | S4 | 2 | 44 | 2x151 | 3340 | 0.3 | 69.2 | 87.8 | 91.1 |

**Table S4: Quality metrics of aligned WGS reads from Illumina and KAPA libraries prepared from manufactured DBS.** Abbreviations: DBS: dried blood spot; CD: coding domain; OMIM: Mendelian inheritance in Man; MT: mitochondrial; SNV: single nucleotide variant; indel: insertion-deletion oligonucleotide variant; CNV: copy number variant.

| **Library Prep** | **Sample ID** | **Raw Yield (Gb)** | **% Reads Align-ed** | **% Duplicate Reads** | **Mean Insert Size (nt)** | **Average Genome Coverage** | **% MIM genes with ≥10X coverage of 100% of CD** | **Coding Domain Variants** | **SNVs** | **Indels** | **CNVs Overlapping MIM gene coding domains** | **Mitoch-ondrial genome coverage** | **Trans-ition/ Trans-version ratio** | **C>T+ A>G/ T>C+ G>A SNP variant ratio** | **Pro-port-ion-ate GC bias** | **Total Length of Mapp-able Genome (Gb)** |
| --- | --- | --- | --- | --- | --- | --- | --- | --- | --- | --- | --- | --- | --- | --- | --- | --- |
| Illumina | 1 | 167 | 97.7% | 9.4% | 466 | 45.2 | 98.2% | 24,499 | 3,976,208 | 940,505 | 3 | 3,419 | 2.03 | 1.04 | -0.09 | ND |
|  | 4 | 151 | 97.8% | 9.0% | 456 | 41.2 | 96.9% | 24,141 | 3,918,968 | 928,905 | 2 | 4,509 | 2.03 | 1.05 | -0.04 | ND |
|  | 10 | 137 | 97.9% | 15.6% | 422 | 36 | 93.9% | 24,989 | 3,826,902 | 951,538 | 18 | 1,714 | 2.03 | 1.05 | -0.07 | ND |
|  | 11_1 | 118 | 97.7% | 15.1% | 472 | 31.2 | 93.2% | 26,115 | 3,970,638 | 979,030 | 13 | 3,167 | 2.04 | 1.05 | -0.07 | 2.67 |
|  | 13_1 | 120 | 97.8% | 8.4% | 429 | 33.1 | 96.9% | 29,540 | 4,768,773 | 1,094,754 | 11 | 2,451 | 2.04 | 1.05 | -0.02 | ND |
|  | 14 | 145 | 97.8% | 15.6% | 412 | 38.1 | 94.1% | 24,942 | 3,838,925 | 955,966 | 12 | 2,194 | 2.03 | 1.05 | -0.02 | ND |
|  | 18 | 128 | 97.7% | 13.0% | 428 | 34.8 | 96.1% | 25,446 | 3,895,926 | 970,452 | 10 | 2,544 | 2.03 | 1.05 | -0.23 | ND |
|  | 19 | 125 | 97.7% | 13.4% | 441 | 33.8 | 92.8% | 25,342 | 3,835,352 | 954,747 | 14 | 2,166 | 2.03 | 1.05 | -0.14 | ND |
|  | 20 | 155 | 97.8% | 13.7% | 356 | 41.6 | 95.4% | 25,117 | 3,831,970 | 957,613 | 11 | 2,928 | 2.03 | 1.04 | 0.00 | ND |
|  | 21 | 159 | 98.1% | 13.2% | 368 | 43.3 | 97.3% | 24,961 | 3,844,408 | 961,100 | 10 | 8,310 | 2.03 | 1.04 | 0.01 | 2.67 |
|  | 22 | 149 | 98.0% | 13.5% | 378 | 40.3 | 94.8% | 24,823 | 3,803,526 | 948,193 | 16 | 3,778 | 2.03 | 1.04 | -0.16 | ND |
|  | 23 | 140 | 98.1% | 9.1% | 367 | 39.7 | 93.6% | 25,286 | 3,913,982 | 975,878 | 20 | 3,870 | 2.03 | 1.05 | 0.13 | ND |
|  | 24 | 135 | 97.9% | 14.5% | 386 | 36 | 92.3% | 25,249 | 3,861,366 | 959,910 | 7 | 3,059 | 2.03 | 1.05 | 0.10 | ND |
|  | 25 | 153 | 98.0% | 13.4% | 376 | 41.4 | 96.0% | 25,258 | 3,828,696 | 958,608 | 10 | 6,523 | 2.03 | 1.05 | 0.09 | ND |
|  | 11_2 | 146 | 97.9% | 12.8% | 409 | 39.7 | 95.7% | 25,991 | 3,968,906 | 986,333 | 14 | 4,811 | 2.03 | 1.04 | -0.31 | 2.67 |
|  | 13_2 | 140 | 97.9% | 8.6% | 400 | 40 | 96.0% | 25,074 | 3,831,223 | 957,676 | 15 | 2,527 | 2.03 | 1.05 | -0.05 | ND |
|  | 1 | 151 | 97.7% | 8.5% | 482 | 41.5 | 98.3% | 24,533 | 3,974,243 | 937,622 | 4 | 2,675 | 2.03 | 1.04 | -0.09 | ND |
|  | 4 | 124 | 97.2% | 8.7% | 465 | 33.6 | 94.9% | 24,156 | 3,908,774 | 914,642 | 2 | 2,675 | 2.03 | 1.04 | -0.18 | ND |
|  | 10 | 131 | 98.0% | 15.3% | 437 | 34.6 | 92.5% | 25,004 | 3,828,786 | 950,945 | 45 | 777 | 2.03 | 1.04 | -0.13 | ND |
|  | 11_1 | 144 | 97.8% | 13.6% | 442 | 38.7 | 95.6% | 26,013 | 3,971,201 | 986,218 | 12 | 4,139 | 2.03 | 1.04 | -0.07 | ND |
|  | 13_1 | 124 | 97.8% | 15.7% | 444 | 32.6 | 96.3% | 30,435 | 4,670,651 | 1,135,905 | 16 | 3,666 | 2.03 | 1.05 | 0.08 | 2.67 |
|  | 14 | 127 | 97.9% | 14.7% | 435 | 34 | 93.0% | 25,023 | 3,841,870 | 953,906 | 10 | 4,171 | 2.03 | 1.05 | 0.10 | 2.67 |
|  | 18 | 117 | 97.7% | 13.0% | 444 | 31.7 | 91.9% | 25,417 | 3,897,828 | 968,605 | 11 | 4,014 | 2.03 | 1.05 | 0.01 | 2.67 |
|  | 19 | 151 | 97.6% | 14.9% | 442 | 40 | 95.2% | 25,252 | 3,831,281 | 955,401 | 10 | 2,657 | 2.03 | 1.05 | 0.06 | 2.67 |
| Avg. |  | 139 | 97.8% | 12.6% | 423 | 37.6 | 95.0% | 25,525 | 3,951,683 | 970,186 | 12 | 3,448 | 2.03 | 1.05 | -0.04 | 2.67 |
| SD |  | 14 | 0.2% | 2.6% | 36.1 | 4.0 | 1.9% | 1,467 | 243,712 | 48,051 | 8.42 | 1,561 | 0.00 | 0.00 | 0.11 | 0.00 |
| Median |  | 140 | 97.8% | 13.4% | 432 | 38.4 | 95.3% | 25,183 | 3,878,646 | 957,645 | 11 | 3,113 | 2.03 | 1.05 | -0.05 | 2.67 |
| KAPA | 1 | 150 | 98.5% | 10.2% | 355 | 42.2 | 90.8% | 24,624 | 3,796,486 | 924,123 | 3399 | 7,161 | 2.04 | **1.04** | 0.78 | 2.67 |
|  | 2 | 134 | 98.4% | 10.5% | 322 | 37.7 | 96.9% | 30,061 | 4,629,653 | 1,109,978 | 19 | 3,053 | 2.04 | **1.05** | 0.14 | 2.67 |
|  | 3 | 153 | 98.1% | 11.6% | 166 | 42.3 | 92.0% | 24,760 | 3,809,095 | 925,033 | 7 | 45,617 | 2.01 | 1.05 | 0.05 | 2.67 |
|  | 4 | 141 | 98.7% | 11.0% | 338 | 37.7 | 87.1% | 23,460 | 3,877,648 | 907,633 | 802 | 3,414 | 2.04 | 1.05 | -0.14 | 2.67 |
|  | 5 | 159 | 98.2% | 9.7% | 340 | 44.9 | 94.9% | 25,642 | 3,876,005 | 952,271 | 9 | 6,585 | 2.02 | 1.05 | -0.16 | 2.67 |
|  | 6 | 138 | 98.3% | 10.0% | 308 | 38.8 | 96.9% | 24,775 | 3,823,390 | 933,632 | 19 | 3,686 | 2.04 | 1.05 | -0.02 | 2.66 |
|  | 7 | 139 | 98.2% | 10.7% | 324 | 38.7 | 94.5% | 24,950 | 3,793,367 | 929,337 | 57 | 4,355 | 2.04 | 1.04 | 0.15 | 2.67 |
|  | 8 | 126 | 98.4% | 10.9% | 312 | 35.3 | 95.9% | 25,077 | 3,833,349 | 937,397 | 5 | 4,252 | 2.03 | 1.05 | 0.08 | 2.66 |
|  | 9 | 222 | 98.3% | 10.7% | 327 | 59.1 | 97.5% | 24,674 | 3,910,898 | 922,371 | 11 | 8,494 | 2.04 | 1.05 | -0.09 | 2.66 |
|  | 10 | 123 | 98.2% | 9.3% | 329 | 34.9 | 93.5% | 24,920 | 3,788,664 | 927,058 | 19 | 4,514 | 2.01 | 1.05 | -0.06 | 2.67 |
|  | 11 | 159 | 98.4% | 11.3% | 374 | 44.1 | 95.5% | 28,137 | 4,313,297 | 1,041,608 | 324 | 8,045 | 2.02 | 1.05 | -0.19 | 2.67 |
|  | 12 | 126 | 98.3% | 10.1% | 338 | 35.4 | 93.5% | 24,826 | 3,799,191 | 930,170 | 95 | 3,837 | 2.04 | 1.05 | 0.17 | 2.67 |
|  | 13 | 168 | 98.7% | 13.4% | 477 | 45.9 | 96.5% | 25,306 | 3,858,340 | 1,000,164 | 122 | 2,809 | 1.98 | 1.04 | 0.13 | 2.67 |
|  | 14 | 128 | 98.3% | 10.7% | 277 | 33.7 | 90.8% | 23,914 | 3,875,802 | 889,363 | 24 | 5,517 | 2.01 | 1.05 | 0.03 | 2.67 |
|  | 15 | 191 | 98.0% | 12.6% | 360 | 49.4 | 96.6% | 24,262 | 3,908,065 | 910,914 | 8 | 7,107 | 2.03 | 1.05 | -0.06 | 2.67 |
|  | 16 | 124 | 98.2% | 10.7% | 267 | 34.7 | 92.8% | 30,024 | 4,620,648 | 1,106,103 | 24 | 4,692 | 2.04 | 1.05 | 0.04 | 2.66 |
|  | 17 | 151 | 98.2% | 11.1% | 176 | 42.1 | 95.9% | 29,805 | 4,597,642 | 1,093,977 | 15 | 5,208 | 2.04 | 1.05 | 0.08 | 2.66 |
|  | 1 | 186 | 98.6% | 11.9% | 392 | 49.1 | 93.2% | 24,192 | 3,890,452 | 918,085 | 455 | 4,377 | 2.02 | 1.05 | -0.07 | 2.67 |
|  | 2 | 152 | 98.6% | 10.8% | 291 | 40.3 | 94.4% | 24,278 | 3,885,525 | 899,205 | 11 | 2,763 | 2.02 | 1.05 | -0.16 | 2.67 |
|  | 3 | 164 | 98.3% | 10.0% | 366 | 44 | 80.3% | 24,334 | 3,925,503 | 920,647 | 1036 | 41,267 | 2.02 | 1.05 | 0.02 | 2.67 |
|  | 4 | 166 | 98.4% | 10.7% | 282 | 44 | 95.6% | 24,676 | 3,930,421 | 914,413 | 6 | 30,749 | 2.04 | 1.04 | -0.04 | 2.67 |
|  | 5 | 151 | 98.2% | 9.7% | 258 | 40.5 | 94.9% | 25,224 | 4,028,693 | 935,185 | 527 | 6,741 | 2.03 | 1.05 | 0.56 | 2.67 |
|  | 6 | 175 | 98.6% | 11.4% | 379 | 46.6 | 85.4% | 24,393 | 3,962,043 | 931,888 | 868 | 11,552 | 2.03 | 1.05 | -0.12 | 2.67 |
|  | 7 | 133 | 98.4% | 10.5% | 301 | 35.4 | 97.1% | 24,324 | 3,885,962 | 904,657 | 2 | 3,610 | 2.03 | 1.05 | -0.25 | 2.67 |
|  | 8 | 246 | 98.2% | 12.5% | 287 | 63.7 | 97.8% | 23,857 | 3,905,492 | 921,577 | 12 | 6,436 | 2.03 | 1.05 | 0.11 | 2.67 |
|  | 9 | 251 | 97.7% | 11.8% | 179 | 64.8 | 97.2% | 24,737 | 3,913,059 | 916,348 | 2 | 10,632 | 2.03 | 1.04 | 0.25 | 2.66 |
|  | 10 | 190 | 98.2% | 11.9% | 314 | 49.6 | 94.8% | 25,057 | 3,967,099 | 933,370 | 66 | 6,353 | 2.02 | 1.04 | -0.09 | 2.67 |
|  | 11 | 161 | 97.8% | 12.3% | 285 | 44 | 94.4% | 25,709 | 3,866,656 | 944,885 | 4 | 5,388 | 2.04 | 1.05 | 0.35 | 2.67 |
|  | 12 | 183 | 98.5% | 11.7% | 300 | 48 | 97.4% | 24,313 | 3,922,361 | 919,166 | 32 | 4,345 | 2.03 | 1.05 | 0.15 | 2.67 |
|  | 13 | 142 | 98.8% | 11.0% | 302 | 37.9 | 97.0% | 24,376 | 3,920,145 | 903,814 | 7 | 3,126 | 2.04 | 1.04 | -0.04 | 2.67 |
|  | 14 | 143 | 98.2% | 9.3% | 207 | 38.3 | 94.2% | 24,416 | 3,877,255 | 899,806 | 351 | 4,925 | 2.03 | 1.05 | 0.32 | 2.66 |
|  | 15 | 150 | 98.3% | 11.1% | 278 | 39.4 | 96.9% | 24,344 | 3,912,332 | 910,758 | 61 | 4,403 | 2.03 | 1.05 | 0.06 | 2.67 |
|  | 16 | 177 | 98.0% | 10.5% | 228 | 46.7 | 97.2% | 24,498 | 3,917,575 | 906,083 | 3 | 5,104 | 2.03 | 1.05 | 0.03 | 2.67 |
|  | 17 | 127 | 98.1% | 10.6% | 321 | 33.6 | 81.2% | 24,233 | 3,873,067 | 888,270 | 159 | 3,444 | 2.02 | 1.05 | -0.03 | 2.67 |
|  | 1 | 160 | 97.7% | 10.3% | 247 | 42 | 95.1% | 24,564 | 3,891,727 | 901,304 | 5 | 4,724 | 2.03 | 1.05 | -0.19 | 2.67 |
|  | 2 | 158 | 98.6% | 11.5% | 376 | 41.9 | 94.2% | 24,228 | 3,891,372 | 912,330 | 145 | 5,812 | 2.02 | 1.05 | 0.08 | 2.67 |
|  | 3 | 159 | 98.4% | 10.6% | 384 | 44.6 | 90.5% | 24,832 | 3,779,735 | 925,516 | 271 | 5,524 | 2.04 | 1.05 | 0.23 | 2.67 |
|  | 5 | 220 | 98.2% | 12.2% | 196 | 57.2 | 97.3% | 27,760 | 4,426,478 | 1,027,196 | 8 | 9,986 | 2.03 | 1.05 | 0.00 | 2.67 |
|  | 6 | 135 | 98.3% | 11.0% | 373 | 35.9 | 81.3% | 24,060 | 3,885,219 | 909,090 | 249 | 4,920 | 2.02 | 1.05 | 0.02 | 2.67 |
| Avg. |  | 160 | 98.3% | 11.0% | 306 | 43.2 | 93.6% | 25,170 | 3,963,326 | 940,634 | 237 | 8,065 | 2.03 | 1.05 | 0.05 | 2.67 |
| SD |  | 32 | 0.3% | 0.9% | 66.3 | 7.7 | 4.6% | 1,654 | 226,097 | 57,237 | 580 | 9,504 | 0.01 | 0.00 | 0.20 | 0.00 |
| Median |  | 153 | 98.3% | 10.8% | 312 | 42.1 | 94.9% | 24,674 | 3,891,372 | 922,371 | 24 | 5,104 | 2.03 | 1.05 | 0.03 | 2.67 |
| KAPA | 1 | 200.3 | 98.7% | 19.20% | 409.0 | 48.5 | 96.84% | 24,346 | 3,916,830 | 916,832 | 3 | nd | 2.03 | 1.047 | -0.07 | 2.67 |
|  | 2 | 163.0 | 98.4% | 14.80% | 394.8 | 41.4 | 95.97% | 25,187 | 3,975,940 | 923,041 | 4 | nd | 2.03 | 1.048 | 0.04 | 2.67 |
|  | 3 | 136.5 | 98.8% | 9.60% | 408.8 | 37.1 | 97.69% | 24,352 | 3,933,824 | 896,828 | 14 | nd | 2.02 | 1.048 | 0.12 | 2.67 |
|  | 4 | 161.0 | 98.6% | 11.00% | 381.3 | 43.0 | 96.92% | 24,659 | 3,908,034 | 910,782 | 31 | nd | 2.03 | 1.047 | 0.18 | 2.67 |
|  | 5 | 189.7 | 98.8% | 10.50% | 382.3 | 51.2 | 98.36% | 24,718 | 3,946,106 | 929,291 | 16 | nd | 2.02 | 1.047 | 0.17 | 2.67 |
|  | 6 | 142.1 | 98.5% | 8.60% | 415.6 | 38.8 | 96.38% | 24,658 | 3,903,231 | 901,676 | 9 | nd | 2.03 | 1.047 | 0.15 | 2.67 |
|  | 7 | 190.5 | 98.7% | 12.70% | 400.1 | 49.9 | 97.83% | 24,669 | 3,914,092 | 917,267 | 95 | nd | 2.02 | 1.047 | 0.24 | 2.67 |
|  | 8 | 170.7 | 98.7% | 10.60% | 395.6 | 45.8 | 97.20% | 27,828 | 4,449,910 | 1,029,642 | 10 | nd | 2.03 | 1.053 | 0.12 | 2.67 |
|  | 9 | 163.6 | 98.6% | 10.70% | 386.4 | 43.8 | 97.05% | 24,434 | 3,909,694 | 921,106 | 19 | nd | 2.02 | 1.042 | 0.18 | 2.67 |
|  | 10 | 173.0 | 98.7% | 9.50% | 415.1 | 47.1 | 98.14% | 29,575 | 4,746,463 | 1,081,171 | 13 | nd | 2.03 | 1.055 | 0.08 | 2.67 |
|  | 11 | 179.8 | 98.7% | 13.60% | 415.8 | 46.7 | 97.78% | 24,705 | 3,934,453 | 916,316 | 4 | nd | 2.01 | 1.047 | 0.07 | 2.67 |
|  | 12 | 174.6 | 98.7% | 12.30% | 299.5 | 45.9 | 97.15% | 24,823 | 3,941,611 | 920,065 | 10 | nd | 2.02 | 1.048 | 0.08 | 2.67 |
|  | 13 | 194.3 | 98.5% | 14.60% | 391.2 | 49.5 | 97.66% | 25,483 | 4,056,403 | 947,695 | 17 | nd | 2.03 | 1.048 | 0.11 | 2.67 |
|  | 14 | 240.5 | 98.8% | 10.40% | 308.0 | 64.9 | 98.31% | 24,711 | 3,980,954 | 940,746 | 7 | nd | 2.03 | 1.046 | -0.02 | 2.67 |
|  | 15 | 178.5 | 98.6% | 9.80% | 454.6 | 48.3 | 97.92% | 24,581 | 3,912,119 | 903,558 | 3 | nd | 2.02 | 1.046 | 0.02 | 2.67 |
|  | 16 | 186.7 | 98.5% | 9.90% | 468.8 | 50.4 | 97.94% | 24,030 | 3,911,614 | 901,162 | 12 | nd | 2.02 | 1.046 | 0.04 | 2.67 |
|  | 17 | 200.9 | 98.7% | 13.30% | 452.4 | 52.3 | 97.92% | 25,009 | 3,948,859 | 926,984 | 7 | nd | 2.02 | 1.050 | 0.14 | 2.67 |
|  | 18 | 150.2 | 98.7% | 9.00% | 430.7 | 41.1 | 97.96% | 24,274 | 3,934,929 | 903,469 | 2 | nd | 2.03 | 1.046 | 0.00 | 2.67 |
|  | 19 | 148.5 | 98.4% | 9.20% | 418.9 | 40.3 | 95.91% | 24,024 | 3,874,838 | 888,758 | 7 | nd | 2.03 | 1.047 | 0.07 | 2.67 |
|  | 20 | 178.8 | 98.2% | 11.80% | 432.0 | 46.9 | 97.21% | 24,427 | 3,906,648 | 902,079 | 8 | nd | 2.03 | 1.046 | -0.09 | 2.67 |
|  | 21 | 143.6 | 98.6% | 11.20% | 443.1 | 38.2 | 97.67% | 24,259 | 3,907,928 | 893,522 | 4 | nd | 2.03 | 1.046 | 0.05 | 2.67 |
|  | 22 | 205.0 | 98.5% | 8.40% | 458.7 | 56.2 | 98.25% | 24,247 | 3,883,571 | 906,849 | 9 | nd | 2.02 | 1.047 | -0.01 | 2.67 |
|  | 23 | 154.5 | 98.5% | 7.10% | 412.4 | 43.0 | 97.79% | 24,300 | 3,978,708 | 913,144 | 17 | nd | 2.03 | 1.046 | 0.02 | 2.67 |
|  | 24 | 133.9 | 98.5% | 8.70% | 435.8 | 36.7 | 94.87% | 24,339 | 3,924,305 | 890,949 | 26 | nd | 2.03 | 1.049 | 0.00 | 2.67 |
|  | 25 | 170.7 | 98.5% | 8.70% | 386.1 | 46.6 | 97.75% | 24,501 | 3,908,433 | 906,768 | 5 | nd | 2.03 | 1.049 | 0.00 | 2.67 |
| Avg. |  | 173.2 | 98.6% | 11.01% | 407.9 | 46.1 | 97.38% | 24,886 | 3,984,380 | 923,588 | 14 | nd | 2.03 | 1.047 | 0.07 | 2.67 |
| Median |  | 173.0 | 98.6% | 10.50% | 412.4 | 46.6 | 97.69% | 24,581 | 3,924,305 | 913,144 | 9 | nd | 2.03 | 1.047 | 0.07 | 2.67 |

**Table S5: Table S5: Quality control metrics for secondary analysis of WGS from DBS and blood.** Abbreviations: DBS: dried blood spot; CD: coding domain; MIM: Mendelian inheritance in Man; SNV: single nucleotide variant; indel: insertion-deletion oligonucleotide variant; CNV: copy number variant; Avg.: Average; Med.: Median; Cov.: Coverage.

| **Method** | **QC metrics** | **Raw Yield (Gb)** | **Reads Map-ped** | **Dupli- cate Reads** | **Align-ed Yield (Gb)** | **Mean insert size (nt)** | **Avg. Cov.** | **Avg. Cov. of MIM genes** | **% MIM genes with 100% CD Cov. ≥10X** | **Total coding variants** | **Total SNVs** | **Total Indels** | **CNVs Overlap-ping MIM gene CDs** | **Mito-chond-rial Genome Cov.** | **Sample size** |
| --- | --- | --- | --- | --- | --- | --- | --- | --- | --- | --- | --- | --- | --- | --- | --- |
| DBS Illumina | Avg. | 139.0 | 97.8% | 12.6% | 136.0 | 423 | 37.6 | 36.0 | 95.03% | 25,525 | 3,951,683 | 970186 | 12 | 3448 | 24 |
|  | Med. | 140.1 | 97.8% | 13.4% | 137.3 | 432 | 38.4 | 35.3 | 95.31% | 25,183 | 3,878,646 | 957645 | 11 | 3113 | 24 |
| DBS KAPA | Avg. | 160.5 | 98.3% | 11.0% | 157.7 | 306 | 43.2 | 42.6 | 93.56% | 25,170 | 3,963,326 | 940634 | 237 | 8197 | 39 |
|  | Med. | 153.1 | 98.3% | 10.8% | 150.2 | 312 | 42.1 | 41.5 | 94.87% | 24,674 | 3,891,372 | 922371 | 24 | 5156 | 39 |
| Blood Illumina | Avg. | 147.9 | 99.0% | 11.8% | 145.8 | 422 | 41.0 | 41.1 | 95.39% | 25,255 | 3,862,520 | 948,453 | 23 | 10598 | 24 |
|  | Med. | 142.7 | 99.1% | 11.9% | 141.3 | 424 | 40.2 | 40.9 | 95.62% | 25,054 | 3,834,070 | 942,348 | 11 | 10562 | 24 |
| Blood KAPA | Avg. | 160.9 | 98.6% | 10.8% | 158.6 | 383 | 43.0 | 45.0 | 96.63% | 25,078 | 3,996,783 | 931,744 | 22 | 10820 | 48 |
|  | Med. | 157.4 | 98.6% | 10.8% | 154.1 | 387 | 41.8 | 42.4 | 97.39% | 24,800 | 3,935,857 | 920,021 | 8 | 5152 | 48 |

**Table S6: Quality metrics of aligned WGS reads from Illumina libraries prepared from California State Biobank DBS.** Abbreviations: DBS: dried blood spot; OMIM: Mendelian inheritance in Man; Nk: not known.

| **ID** | **Year Collected** | **DBS Paper Type** | **# of Punches avail-able** | **DNA yield (ng)** | **A260/ A280 ratio** | **Library input DNA (ng)** | **Library yield (nM)** | **Raw WGS Yield (GB)** | **% Reads Map-ped** | **% Dupli-cate Reads** | **Mean Insert Size (bp)** | **Avg. Genome Cover-age** | **Avg. Cover-age of MIM genes** | **% MIM genes with >10X cover-age of 100% coding domain** | **Prop-ortion-ate GC bias** |
| --- | --- | --- | --- | --- | --- | --- | --- | --- | --- | --- | --- | --- | --- | --- | --- |
| 1 | 2020 | Whatman | 10 | 544 | 1.77 | 425 | 8.0 | 155 | 99.1% | 12.9% | 433 | 42.8 | 40.3 | 97.2% | 0.01 |
| 2 | 2020 | Whatman | 10 | 689 | 1.81 | 500 | 7.8 | 123 | 99.1% | 11.4% | 437 | 34.3 | 32.4 | 95.0% | -0.03 |
| 3 | 2020 | Whatman | 10 | 405 | 1.6 | 316 | 10.3 | 108 | 99.3% | 8.3% | 280 | 31.4 | 30.6 | 92.4% | -0.06 |
| 4 | 2020 | Whatman | 12 | 621 | 1.67 | 485 | 10.0 | 130 | 99.3% | 8.4% | 443 | 37.6 | 37.7 | 97.6% | 0.10 |
| 5 | 2019 | Whatman | 8 | 622 | 1.64 | 486 | 7.0 | 158 | 99.3% | 8.9% | 443 | 45.6 | 44.9 | 97.0% | 0.10 |
| 6 | 2018 | Whatman | 12 | 638 | 1.73 | 499 | 13.5 | 114 | 99.4% | 8.7% | 406 | 33 | 32.4 | 93.6% | 0.09 |
| 7 | 2015 | Whatman | 12 | 331 | 1.83 | 258 | 13.1 | 97 | 99.4% | 8.4% | 249 | 28 | 29.5 | 90.8% | 0.07 |
| 8 | 2015 | Whatman | 8 | 343 | 1.7 | 268 | 6.8 | 149 | 99.3% | 8.4% | 337 | 43.3 | 42.3 | 97.6% | -0.16 |
| 9 | 2014 | Whatman | 12 | 459 | 1.7 | 359 | 10.2 | 142 | 99.3% | 7.8% | 436 | 41.5 | 41.2 | 96.0% | 0.03 |
| 10 | 2014 | Whatman | 12 | 590 | 1.87 | 461 | 10.7 | 101 | 99.4% | 9.3% | 430 | 29 | 26 | 75.1% | -0.21 |
| 11 | 2013 | PerkinElmer | 12 | 697 | 1.71 | 545 | 11.2 | 195 | 99.3% | 8.4% | 453 | 56.6 | 56.7 | 98.0% | 0.13 |
| 12 | 2013 | PerkinElmer | 12 | 934 | 1.73 | 500 | 12.2 | 158 | 99.0% | 13.8% | 433 | 42.9 | 39.6 | 95.2% | 0.03 |
| 13 | 2013 | PerkinElmer | 10 | 550 | 1.82 | 430 | 8.0 | 97 | 99.4% | 9.4% | 427 | 27.8 | 25 | 71.4% | -0.19 |
| 14 | 2013 | PerkinElmer | 10 | 769 | 2.11 | 500 | 6.3 | 133 | 99.3% | 7.7% | 462 | 38.7 | 37.9 | 95.5% | 0.02 |
| 15 | 2011 | Whatman | 10 | 606 | 1.66 | 474 | 7.0 | 115 | 99.3% | 8.0% | 427 | 33.6 | 29.3 | 78.5% | -0.37 |
| 16 | 2011 | Whatman | 10 | 582 | 1.72 | 455 | 7.0 | 114 | 99.3% | 8.4% | 420 | 33 | 28.6 | 76.7% | -0.38 |
| 17 | 2007 | Whatman | 6 | 925 | 1.78 | 361 | 7.0 | 143 | 99.1% | 13.5% | 430 | 39 | 36.2 | 94.1% | 0.00 |
| 18 | 2007 | Whatman | 7 | 787 | 1.8 | 500 | 6.6 | 148 | 99.1% | 13.2% | 443 | 40.7 | 38.2 | 94.6% | 0.03 |
| 19 | 2007 | Whatman | 8 | 555 | 1.57 | 434 | 8.2 | 101 | 99.3% | 7.6% | 435 | 29.5 | 25.8 | 69.9% | -0.34 |
| 20 | 2007 | Whatman | 10 | 483 | 1.52 | 378 | 7.9 | 146 | 99.2% | 10.1% | 427 | 41.4 | 38.8 | 95.1% | -0.09 |
| 21 | 2004 | nk | 7 | 343 | 1.63 | 268 | 8.7 | 155 | 99.2% | 10.5% | 411 | 43.8 | 39.5 | 95.3% | -0.16 |
| 22 | 2004 | nk | 8 | 608 | 1.68 | 475 | 10.7 | 98 | 99.3% | 8.0% | 436 | 28.5 | 26.4 | 81.8% | -0.18 |
| 23 | 2004 | nk | 12 | 444 | 1.63 | 347 | 9.8 | 167 | 99.0% | 14.8% | 410 | 45 | 41.4 | 95.5% | -0.01 |
| 24 | 2003 | nk | 10 | 824 | 1.78 | 500 | 8.8 | 134 | 99.3% | 7.3% | 457 | 39.4 | 38.5 | 97.6% | 0.02 |
| 25 | 2003 | nk | 12 | 780 | 1.8 | 500 | 8.3 | 134 | 99.3% | 7.8% | 456 | 39.1 | 37.6 | 97.5% | -0.05 |
| 26 | 2002 | nk | 10 | 846 | 1.72 | 500 | 5.8 | 149 | 99.3% | 8.4% | 447 | 43.3 | 40.8 | 97.7% | -0.04 |
| 27 | 2002 | nk | 12 | 871 | 1.75 | 500 | 8.7 | 149 | 99.3% | 8.7% | 445 | 42.9 | 40.7 | 97.8% | -0.01 |
| 28 | 2000 | nk | 8 | 793 | 1.86 | 500 | 7.0 | 126 | 99.3% | 8.4% | 449 | 36.6 | 35 | 94.6% | -0.04 |
| 29 | 2000 | nk | 12 | 1050 | 1.79 | 500 | 5.8 | 177 | 99.4% | 11.2% | 436 | 49.8 | 43.3 | 96.0% | -0.22 |
| **Avg.** |  |  | **10.1** | **645** | **1.74** | **439** | **8.7** | **135** | **99.3%** | **9.6%** | **421** | **38.6** | **36.4** | **91.6%** | **-0.07** |

**Table S7: Additional quality metrics of aligned WGS reads from Illumina libraries prepared from California State Biobank DBS.** Abbreviations: OMIM: Mendelian inheritance in Man; CNV: copy number variant; CD: Coding Domain.

| **ID** | **Total Variants** | **Total SNVs** | **Total Indels** | **Mitoch-ondrial genome coverage** | **Total CD Variants** | **CNV calls overlap-ping CD of MIM genes** | **Mito-chondrial Variants** | **Average Genome Coverage** | **% Bases >Q30** | **Homo-zygous/ Hetero-zygous SNV Ratio** | **Trans-ition/ Trans-version Ratio** | **C>T+ G>A / T>C+ A>G variant ratio** | **Std. Dev./ Average Genome Coverage** | **Map-pable Genome (Gb)** |
| --- | --- | --- | --- | --- | --- | --- | --- | --- | --- | --- | --- | --- | --- | --- |
| 1 | 4,811,486 | 3,942,078 | 967,418 | 6,567 | 25,657 | 4 | 41 | 40 | 90.5 | 0.62 | 2.03 | 1.05 | 0.19 | 2.67 |
| 2 | 4,811,814 | 3,945,033 | 964,102 | 5,234 | 25,612 | 6 | 41 | 42 | 90.6 | 0.63 | 2.03 | 1.05 | 0.20 | 2.67 |
| 3 | 5,012,050 | 3,895,999 | 966,195 | 2,487 | 27,259 | 12 | 63 | 46 | 89.8 | 0.56 | 2.02 | 1.04 | 0.25 | 2.67 |
| 4 | 5,031,854 | 4,550,087 | 1,113,850 | 2,763 | 27,070 | 9 | 114 | 59 | 89.9 | 0.63 | 2.03 | 1.05 | 0.23 | 2.67 |
| 5 | 5,067,541 | 3,853,063 | 965,110 | 2,941 | 27,648 | 9 | 44 | 42 | 88.5 | 0.59 | 2.03 | 1.04 | 0.22 | 2.67 |
| 6 | 4,998,738 | 4,450,898 | 1,092,034 | 1,842 | 26,979 | 4 | 13 | 45 | 89.2 | 0.59 | 2.03 | 1.05 | 0.24 | 2.67 |
| 7 | 5,575,714 | 4,051,882 | 997,737 | 3,807 | 30,809 | 16 | 84 | 25 | 85.3 | 0.43 | 2.02 | 1.05 | 0.26 | 2.67 |
| 8 | 5,131,708 | 4,060,819 | 1,009,234 | 6,255 | 27,678 | 7 | 42 | 42 | 88.8 | 0.55 | 2.02 | 1.04 | 0.23 | 2.67 |
| 9 | 4,965,571 | 3,965,275 | 986,159 | 2,484 | 27,158 | 7 | 23 | 38 | 87.6 | 0.60 | 2.02 | 1.04 | 0.23 | 2.67 |
| 10 | 4,965,531 | 3,961,741 | 978,682 | 1,995 | 27,144 | 9 | 23 | 39 | 87.8 | 0.60 | 2.03 | 1.05 | 0.26 | 2.67 |
| 11 | 5,000,603 | 3,943,476 | 985,596 | 2,568 | 27,214 | 4 | 13 | 36 | 88.3 | 0.59 | 2.02 | 1.04 | 0.21 | 2.67 |
| 12 | 5,058,793 | 3,898,162 | 957,186 | 5,268 | 27,757 | 9 | 44 | 26 | 85.9 | 0.58 | 2.03 | 1.05 | 0.19 | 2.67 |
| 13 | 5,602,512 | 4,370,367 | 1,066,415 | 1,735 | 30,751 | 15 | 62 | 39 | 88.3 | 0.44 | 2.03 | 1.05 | 0.26 | 2.67 |
| 14 | 5,130,183 | 4,383,797 | 1,081,191 | 2,102 | 27,739 | 8 | 42 | 41 | 88.8 | 0.55 | 2.03 | 1.05 | 0.23 | 2.67 |
| 15 | 5,042,366 | 3,943,288 | 966,061 | 2,498 | 27,175 | 4 | 29 | 34 | 89.1 | 0.58 | 2.03 | 1.04 | 0.27 | 2.67 |
| 16 | 4,965,194 | 3,944,861 | 966,126 | 2,937 | 27,282 | 7 | 81 | 24 | 84.3 | 0.61 | 2.02 | 1.04 | 0.27 | 2.67 |
| 17 | 5,022,892 | 3,768,701 | 929,017 | 4,765 | 27,559 | 11 | 63 | 27 | 85.3 | 0.59 | 2.03 | 1.04 | 0.19 | 2.67 |
| 18 | 5,022,504 | 3,768,344 | 929,351 | 4,595 | 27,695 | 8 | 63 | 27 | 84.9 | 0.59 | 2.03 | 1.04 | 0.19 | 2.67 |
| 19 | 4,974,134 | 3,904,140 | 952,343 | 1,871 | 26,939 | 8 | 29 | 31 | 88.8 | 0.61 | 2.03 | 1.04 | 0.27 | 2.67 |
| 20 | 5,172,245 | 3,913,772 | 970,369 | 2,991 | 28,374 | 9 | 93 | 29 | 87.4 | 0.53 | 2.02 | 1.04 | 0.23 | 2.67 |
| 21 | 5,195,613 | 3,945,342 | 979,211 | 2,881 | 28,666 | 8 | 93 | 40 | 88.3 | 0.53 | 2.02 | 1.04 | 0.23 | 2.67 |
| 22 | 4,930,818 | 3,968,921 | 972,014 | 1,821 | 26,791 | 12 | 41 | 46 | 89.2 | 0.60 | 2.03 | 1.05 | 0.26 | 2.67 |
| 23 | 5,821,702 | 3,926,180 | 963,408 | 6,694 | 31,512 | 15 | 28 | 38 | 88.5 | 0.41 | 2.03 | 1.05 | 0.19 | 2.67 |
| 24 | 5,683,853 | 3,874,132 | 970,125 | 1,540 | 30,978 | 14 | 53 | 33 | 88.0 | 0.44 | 2.02 | 1.04 | 0.23 | 2.67 |
| 25 | 5,058,386 | 3,874,590 | 969,595 | 1,713 | 27,911 | 11 | 29 | 26 | 86.1 | 0.55 | 2.02 | 1.04 | 0.23 | 2.67 |
| 26 | 4,990,619 | 4,002,905 | 997,654 | 1,645 | 27,321 | 7 | 71 | 40 | 88.7 | 0.61 | 2.02 | 1.04 | 0.22 | 2.67 |
| 27 | 5,035,197 | 4,003,526 | 998,144 | 1,654 | 27,280 | 7 | 29 | 41 | 88.3 | 0.62 | 2.02 | 1.04 | 0.22 | 2.67 |
| 28 | 4,974,759 | 3,913,175 | 973,267 | 1,851 | 26,720 | 14 | 114 | 44 | 90.2 | 0.63 | 2.03 | 1.04 | 0.23 | 2.67 |
| 29 | 5,042,366 | 3,908,000 | 978,010 | 2,762 | 27,175 | 4 | 29 | 34 | 89.1 | 0.58 | 2.02 | 1.04 | 0.22 | 2.67 |
| **Avg.** | **5,106,784** | **3,997,674** | **987,779** | **3,113** | **27,788** | **9** | **52** | **37** | **88.1** | **0.57** | **2.03** | **1.04** | **0.23** | **2.67** |
| **SD** | **246,984** | **192,602** | **44,979** | **1,585** | **1,456** | **3.56** | **28.26** | **8.00** | **1.68** | **0.06** | **0.01** | **0.00** | **0.03** | **0.00** |

**Table S8. Concordance analysis of single nucleotide variants (SNVs) and small insertions and deletions (indels) between WGS from EDTA blood and DBS.**

| **Sample ID** | **DBS type** | **Library Preparation Method** | **EDTA Blood Unique SNVs** | **DBS Sample Unique SNVs** | **SNVs Common to Blood and DBS** | **SNV Con-cordance** | **EDTA Blood Unique indels** | **DBS Sample Unique indels** | **indels Common to Blood and DBS** | **indel Con-cordance** |
| --- | --- | --- | --- | --- | --- | --- | --- | --- | --- | --- |
| 1 | FTA | Illumina | 6,634 | 12,514 | 3,041,847 | 99.4% | 4,783 | 7,356 | 492,376 | 97.6% |
| 1 | PC | Illumina | 6,604 | 12,942 | 3,041,883 | 99.4% | 5,146 | 7,517 | 492,001 | 97.5% |
| 1 | FTA | KAPA | 10,553 | 10,186 | 3,037,944 | 99.3% | 7,472 | 6,936 | 489,667 | 97.1% |
| 1 | PC | KAPA | 10,198 | 9,979 | 3,038,268 | 99.3% | 7,648 | 6,731 | 489,520 | 97.1% |
| 1 | PC | KAPA | 17,180 | 9,694 | 3,031,285 | 99.1% | 16,208 | 12,050 | 480,958 | 94.5% |
| 2 | FTA | KAPA | 8,016 | 11,056 | 3,085,526 | 99.4% | 6,285 | 9,108 | 500,429 | 97.0% |
| 2 | PC | KAPA | 10,681 | 10,062 | 3,082,849 | 99.3% | 8,844 | 9,514 | 497,875 | 96.4% |
| 2 | PC | KAPA | 11,154 | 12,799 | 3,082,394 | 99.2% | 7,852 | 9,973 | 498,858 | 96.6% |
| 3 | FTA | KAPA | 14,002 | 13,416 | 3,016,578 | 99.1% | 9,203 | 12,536 | 481,837 | 95.7% |
| 3 | PC | KAPA | 15,203 | 11,353 | 3,015,354 | 99.1% | 10,720 | 12,819 | 480,341 | 95.3% |
| 3 | PC | KAPA | 15,718 | 13,911 | 3,014,836 | 99.0% | 14,005 | 15,152 | 477,055 | 94.2% |
| 4 | FTA | Illumina | 6,608 | 14,026 | 3,029,084 | 99.3% | 5,548 | 8,487 | 489,090 | 97.2% |
| 4 | PC | Illumina | 6,602 | 13,471 | 3,029,090 | 99.3% | 5,229 | 8,281 | 489,405 | 97.3% |
| 4 | FTA | KAPA | 9,172 | 11,351 | 3,026,531 | 99.3% | 6,724 | 8,428 | 487,897 | 97.0% |
| 4 | PC | KAPA | 17,734 | 11,412 | 3,017,943 | 99.0% | 13,233 | 11,790 | 481,408 | 95.1% |
| 5 | FTA | KAPA | 9,350 | 10,497 | 3,025,747 | 99.3% | 7,170 | 7,912 | 488,936 | 97.0% |
| 5 | PC | KAPA | 14,253 | 9,429 | 3,020,826 | 99.2% | 12,816 | 10,299 | 483,308 | 95.4% |
| 5 | PC | KAPA | 15,255 | 8,328 | 3,019,829 | 99.2% | 11,522 | 8,381 | 484,594 | 96.1% |
| 6 | FTA | KAPA | 11,605 | 11,430 | 3,023,977 | 99.2% | 7,768 | 9,358 | 486,238 | 96.6% |
| 6 | PC | KAPA | 18,507 | 11,747 | 3,017,078 | 99.0% | 17,337 | 13,723 | 476,666 | 93.9% |
| 6 | PC | KAPA | 16,898 | 12,726 | 3,018,686 | 99.0% | 12,606 | 11,888 | 481,393 | 95.2% |
| 7 | FTA | KAPA | 11,706 | 12,131 | 3,018,391 | 99.2% | 8,196 | 9,061 | 486,761 | 96.6% |
| 7 | PC | KAPA | 11,982 | 9,901 | 3,018,102 | 99.3% | 9,625 | 7,747 | 485,347 | 96.5% |
| 7 | PC | KAPA | 11,545 | 11,925 | 3,018,549 | 99.2% | 7,153 | 7,457 | 487,808 | 97.1% |
| 8 | FTA | KAPA | 12,171 | 12,580 | 3,456,999 | 99.3% | 8,553 | 9,155 | 561,592 | 96.9% |
| 8 | PC | KAPA | 19,201 | 10,009 | 3,449,945 | 99.2% | 10,866 | 10,124 | 559,300 | 96.4% |
| 9 | FTA | KAPA | 9,668 | 11,421 | 3,036,003 | 99.3% | 7,621 | 8,170 | 493,979 | 96.9% |
| 9 | PC | KAPA | 12,205 | 9,266 | 3,033,444 | 99.3% | 11,382 | 8,782 | 490,227 | 96.0% |
| 10 | FTA | Illumina | 12,423 | 17,305 | 3,682,496 | 99.2% | 15,456 | 15,131 | 594,647 | 95.1% |
| 10 | PC | Illumina | 9,930 | 19,230 | 3,684,996 | 99.2% | 9,013 | 13,653 | 601,095 | 96.4% |
| 10 | FTA | KAPA | 13,685 | 15,381 | 3,681,259 | 99.2% | 10,135 | 12,826 | 599,951 | 96.3% |
| 10 | PC | KAPA | 17,205 | 19,931 | 3,677,745 | 99.0% | 13,125 | 18,942 | 596,933 | 94.9% |
| 10 | PC | KAPA | 24,960 | 17,672 | 3,669,986 | 98.9% | 15,722 | 15,513 | 594,348 | 95.0% |
| 11 | FTA | Illumina | 7,263 | 12,578 | 3,032,422 | 99.3% | 4,653 | 7,046 | 491,035 | 97.7% |
| 11 | FTA | Illumina | 7,634 | 11,911 | 3,032,051 | 99.4% | 5,250 | 7,083 | 490,432 | 97.5% |
| 11 | PC | Illumina | 7,606 | 12,253 | 3,032,083 | 99.3% | 5,361 | 7,053 | 490,312 | 97.5% |
| 11 | PC | KAPA | 13,320 | 9,771 | 3,026,363 | 99.2% | 11,483 | 9,260 | 484,189 | 95.9% |
| 11 | PC | KAPA | 10,029 | 7,838 | 3,029,644 | 99.4% | 6,774 | 5,752 | 488,914 | 97.5% |
| 12 | FTA | KAPA | 19,286 | 11,840 | 3,054,440 | 99.0% | 10,642 | 10,027 | 489,312 | 95.9% |
| 12 | PC | KAPA | 11,758 | 11,809 | 3,061,928 | 99.2% | 10,633 | 10,745 | 489,352 | 95.8% |
| 12 | PC | KAPA | 11,250 | 10,723 | 3,062,431 | 99.3% | 8,289 | 7,456 | 491,701 | 96.9% |
| 13 | FTA | Illumina | 6,939 | 13,588 | 3,140,921 | 99.4% | 5,205 | 7,945 | 509,347 | 97.5% |
| 13 | FTA | Illumina | 7,281 | 14,130 | 3,140,589 | 99.3% | 6,490 | 8,415 | 508,048 | 97.1% |
| 13 | PC | Illumina | 6,661 | 13,913 | 3,141,203 | 99.3% | 5,211 | 7,894 | 509,342 | 97.5% |
| 13 | PC | KAPA | 14,747 | 10,001 | 3,133,111 | 99.2% | 11,227 | 8,275 | 503,324 | 96.3% |
| 14 | FTA | Illumina | 5,721 | 14,666 | 3,060,376 | 99.3% | 4,936 | 6,990 | 497,840 | 97.7% |
| 14 | PC | Illumina | 5,882 | 14,855 | 3,060,207 | 99.3% | 5,311 | 7,091 | 497,473 | 97.6% |
| 14 | PC | KAPA | 8,754 | 9,223 | 3,057,335 | 99.4% | 7,452 | 5,742 | 495,335 | 97.4% |
| 15 | FTA | KAPA | 20,004 | 9,529 | 3,005,792 | 99.0% | 10,573 | 9,175 | 481,854 | 96.1% |
| 15 | PC | KAPA | 16,065 | 9,372 | 3,009,736 | 99.2% | 12,167 | 10,865 | 480,262 | 95.4% |
| 16 | PC | KAPA | 12,941 | 9,208 | 3,006,604 | 99.3% | 6,909 | 8,158 | 483,157 | 97.0% |
| 17 | PC | KAPA | 21,124 | 8,785 | 3,042,819 | 99.0% | 8,869 | 7,140 | 490,338 | 96.8% |
| 18 | FTA | Illumina | 6,575 | 12,035 | 3,026,843 | 99.4% | 7,060 | 9,603 | 484,887 | 96.7% |
| 18 | PC | Illumina | 6,669 | 12,081 | 3,026,750 | 99.4% | 7,521 | 9,621 | 484,420 | 96.6% |
| 19 | FTA | Illumina | 6,308 | 12,797 | 3,018,212 | 99.4% | 7,498 | 10,541 | 483,581 | 96.4% |
| 19 | PC | Illumina | 7,267 | 12,589 | 3,017,251 | 99.3% | 10,097 | 11,305 | 480,977 | 95.7% |
| 20 | FTA | Illumina | 9,123 | 13,756 | 3,039,086 | 99.3% | 5,686 | 9,308 | 490,093 | 97.0% |
| 21 | FTA | Illumina | 9,055 | 15,059 | 3,018,481 | 99.2% | 6,522 | 11,797 | 484,642 | 96.4% |
| 22 | FTA | Illumina | 8,734 | 13,506 | 3,020,837 | 99.3% | 5,178 | 7,724 | 487,306 | 97.4% |
| 23 | FTA | Illumina | 6,892 | 15,744 | 3,059,750 | 99.3% | 6,134 | 11,086 | 493,958 | 96.6% |
| 24 | FTA | Illumina | 9,543 | 17,335 | 3,049,393 | 99.1% | 7,539 | 13,165 | 488,270 | 95.9% |
| 25 | FTA | Illumina | 7,844 | 14,284 | 3,031,351 | 99.3% | 5,393 | 9,119 | 490,201 | 97.1% |

**Supplementary Figure**

**Supplementary Figure 1: Impact of DBS variables on WGS quality. a.** Comparison of age of DBS with genome coverage by WGS of DNA derived from those DBS. **b.** Comparison of age of DBS with average coverage of Mendelian Inheritance in Man genes by WGS of DNA derived from those DBS. **c.** Comparison of age of DBS with raw WGS yield of DNA derived from those DBS. **d.** Comparison of genome coverage by WGS with DBS filter paper type used for California newborn screening.

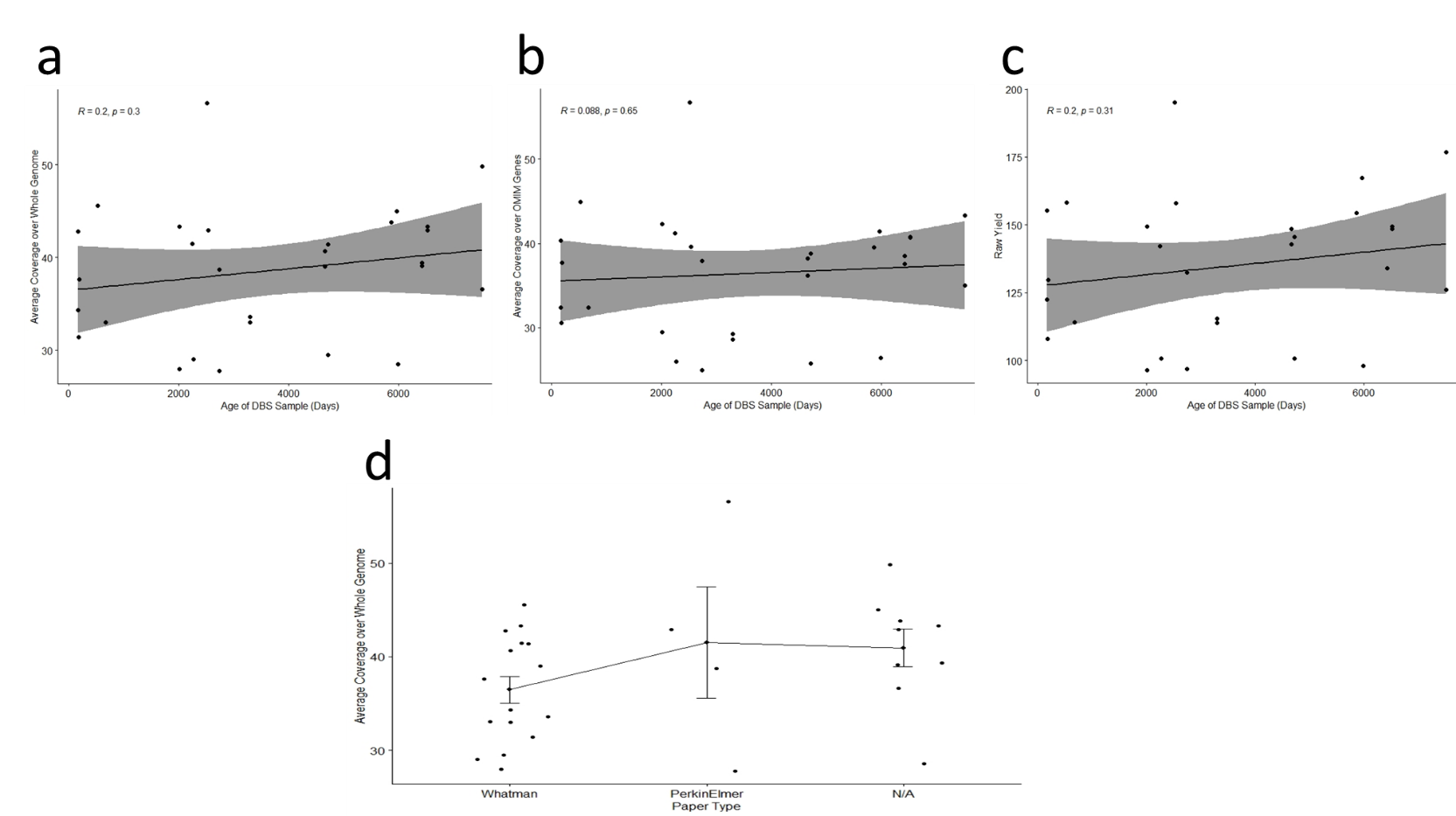

**Figure S2:** Chr 1:187,677,560-187,677,598, an AT-rich, non-coding region showing a heterozygous dinucleotide deletion and a discordant, overlapping heterozygous T>A substitution. Shown, from top to bottom, are the reference nucleotide sequence, average coverage in WGS from 200 unrelated subjects, and coverage and representative reads from a proband (blood sample), father (blood sample), sibling (blood sample), father (DBS, KAPA library) father (DBS, Illumina library), proband (DBS, KAPA library), proband (DBS, Illumina library), and RepeatMasker.

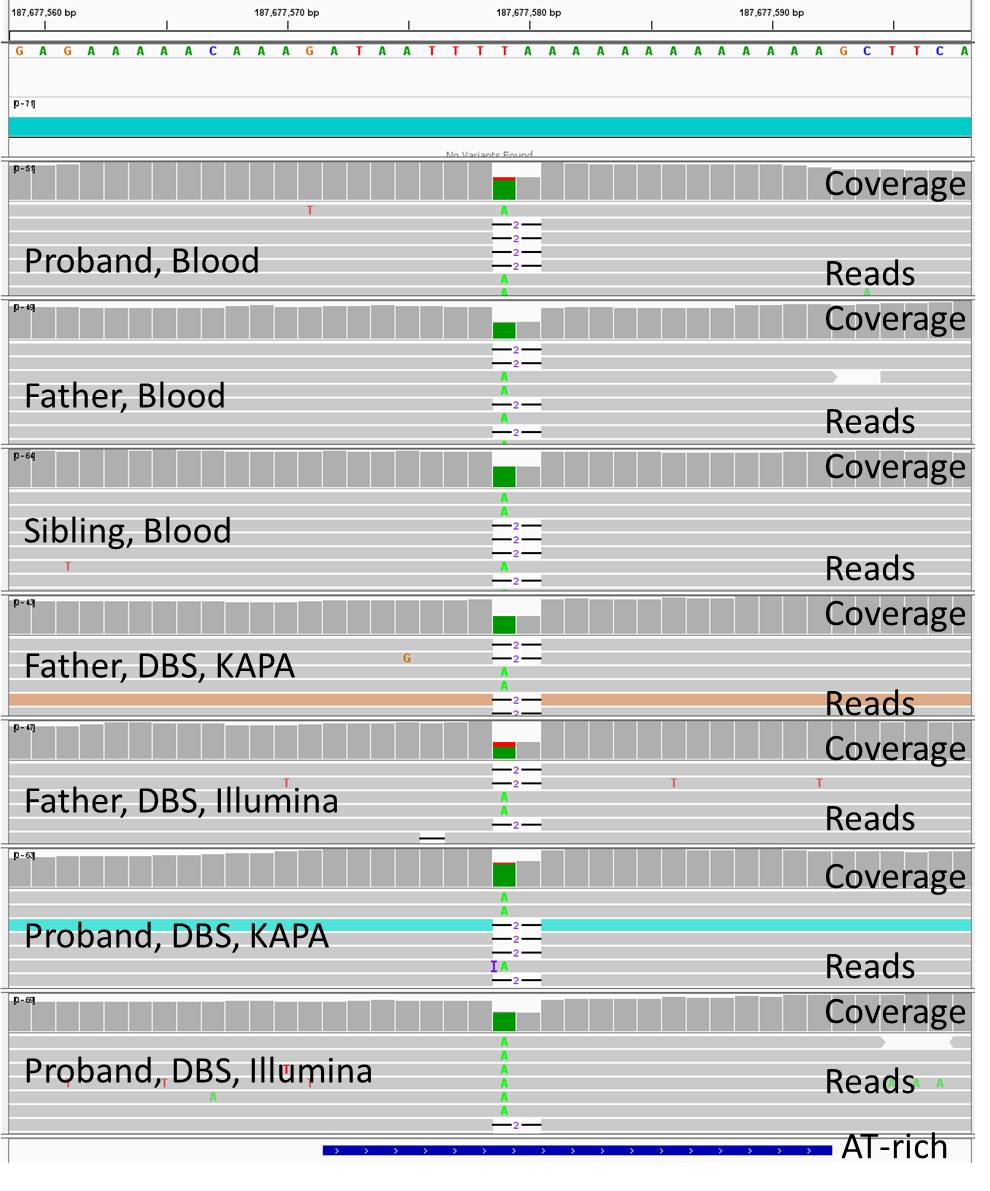

**Figure S3:** Chr 18:21,542,872-21,542,910, a non-coding region featuring an Alu element, showing a heterozygous tetranucleotide deletion and a discordant, overlapping heterozygous A>G substitution. Shown, from top to bottom, are the reference nucleotide sequence, average coverage in WGS from 200 unrelated subjects, and coverage and representative reads from a proband (blood sample), father (blood sample), sibling (blood sample), father (DBS, KAPA library) father (DBS, Illumina library), proband (DBS, KAPA library), proband (DBS, Illumina library), and RepeatMasker.

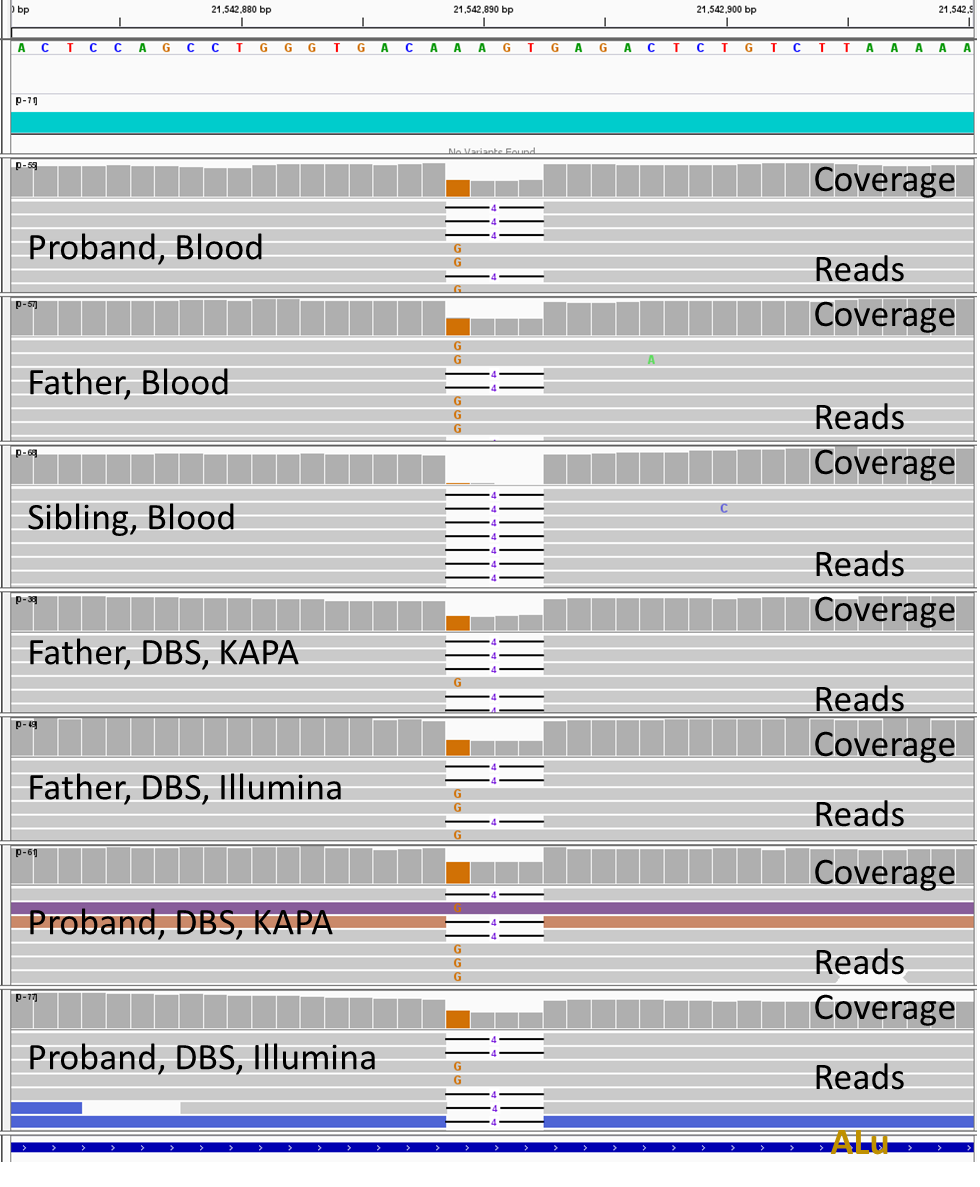
**Figure S4:** Chr 4:171,247,140-171,247,178, an AT-rich, non-coding region showing a heterozygous, single nucleotide deletion with a discordant, overlapping heterozygous T>A substitution in some samples. The sibling is homozygous for the deletion. Shown, from top to bottom, are the reference nucleotide sequence, average coverage in WGS from 200 unrelated subjects, and coverage and representative reads from a proband (blood sample), father (blood sample), sibling (blood sample), father (DBS, KAPA library) father (DBS, Illumina library), proband (DBS, KAPA library), proband (DBS, Illumina library), and RepeatMasker.

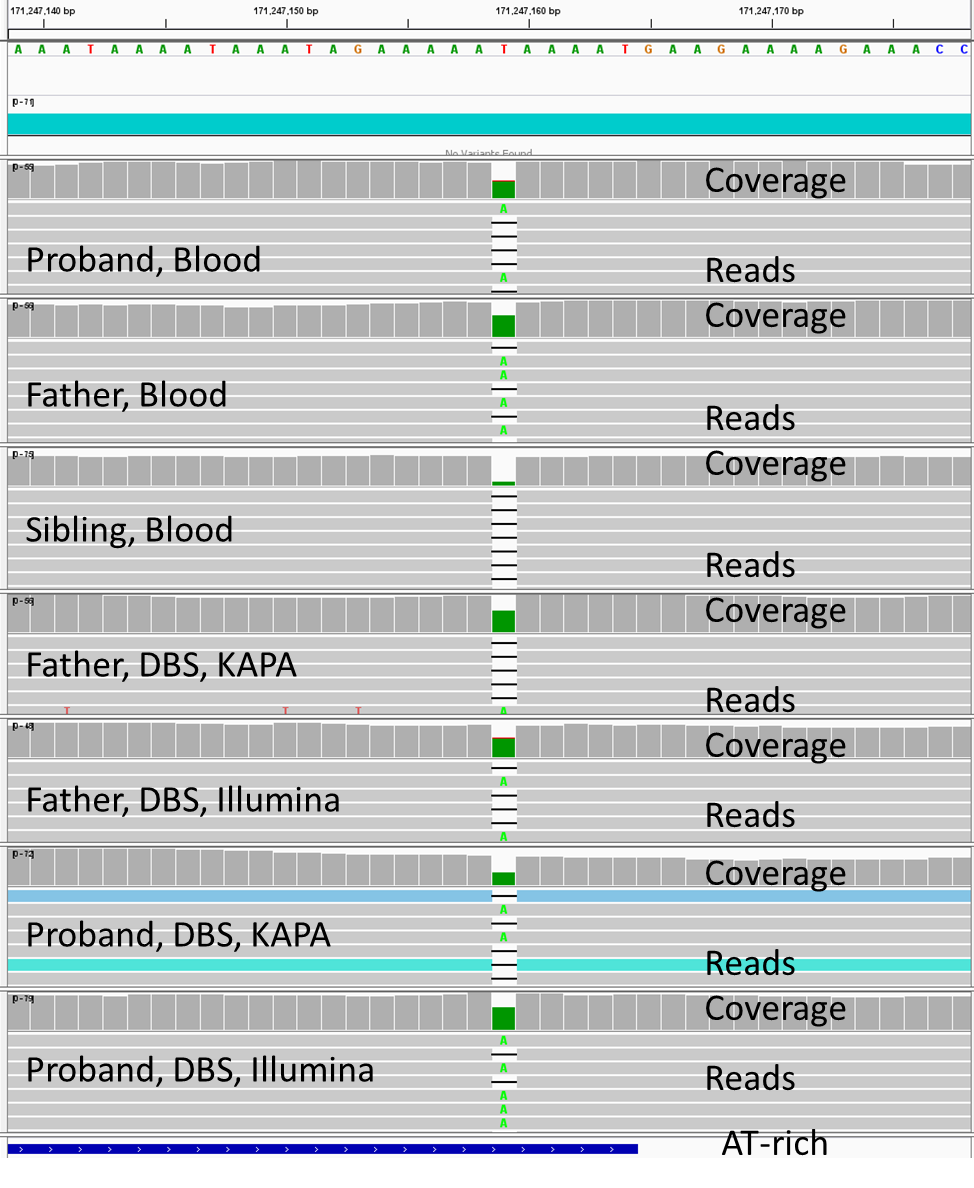

**Figure S5:** Chr 3:5,211,240-5,211,271, a CT-rich region of a LINE1 retrotransposon in an intron of ARL8B showing a heterozygous, single nucleotide deletion with a discordant, overlapping heterozygous C>T substitution in in the father. Shown, from top to bottom, are the reference nucleotide sequence, gene, average WGS coverage of 200 unrelated subjects, and coverage and representative reads from a proband (blood sample), father (blood sample), sibling (blood sample), father (DBS, KAPA library) father (DBS, Illumina library), proband (DBS, KAPA library), proband (DBS, Illumina library), and RepeatMasker.

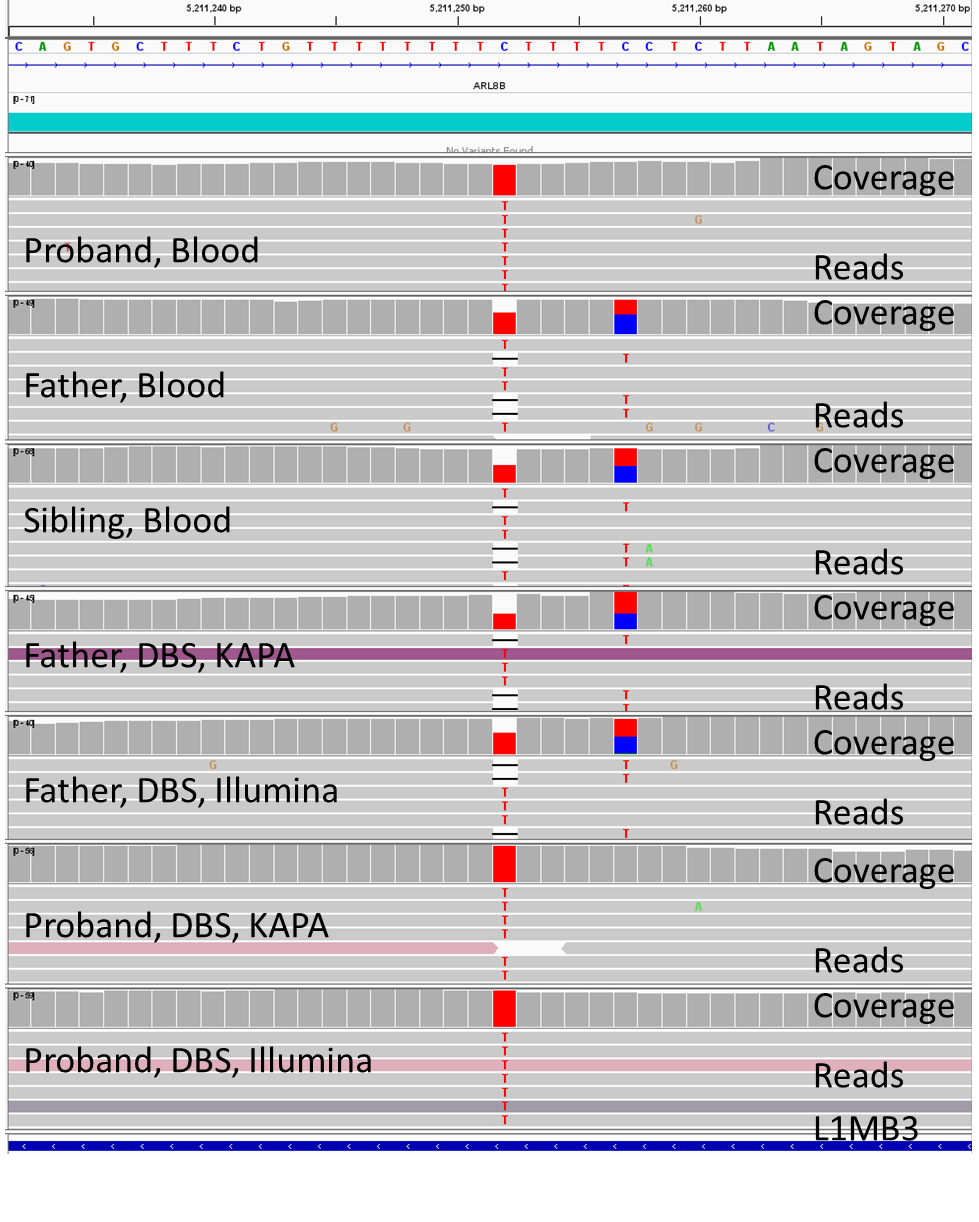

**Figure S6:** Chr 10:132,112,292-132,112,330, a non-coding, TTA-repeat, LINE1-containing region showing 3 discrepant, overlapping variants: A homozygous or heterozygous trinucleotide deletion overlapping with a heterozygous A>T substitution which was also called as a TA>T single nucleotide deletion. This may be a random alignment error. Shown, from top to bottom, are the reference nucleotide sequence, average WGS coverage in 200 unrelated subjects, and coverage and representative reads from a proband (blood sample), father (blood sample), sibling (blood sample), father (DBS, KAPA library) father (DBS, Illumina library), proband (DBS, KAPA library), proband (DBS, Illumina library), and RepeatMasker.

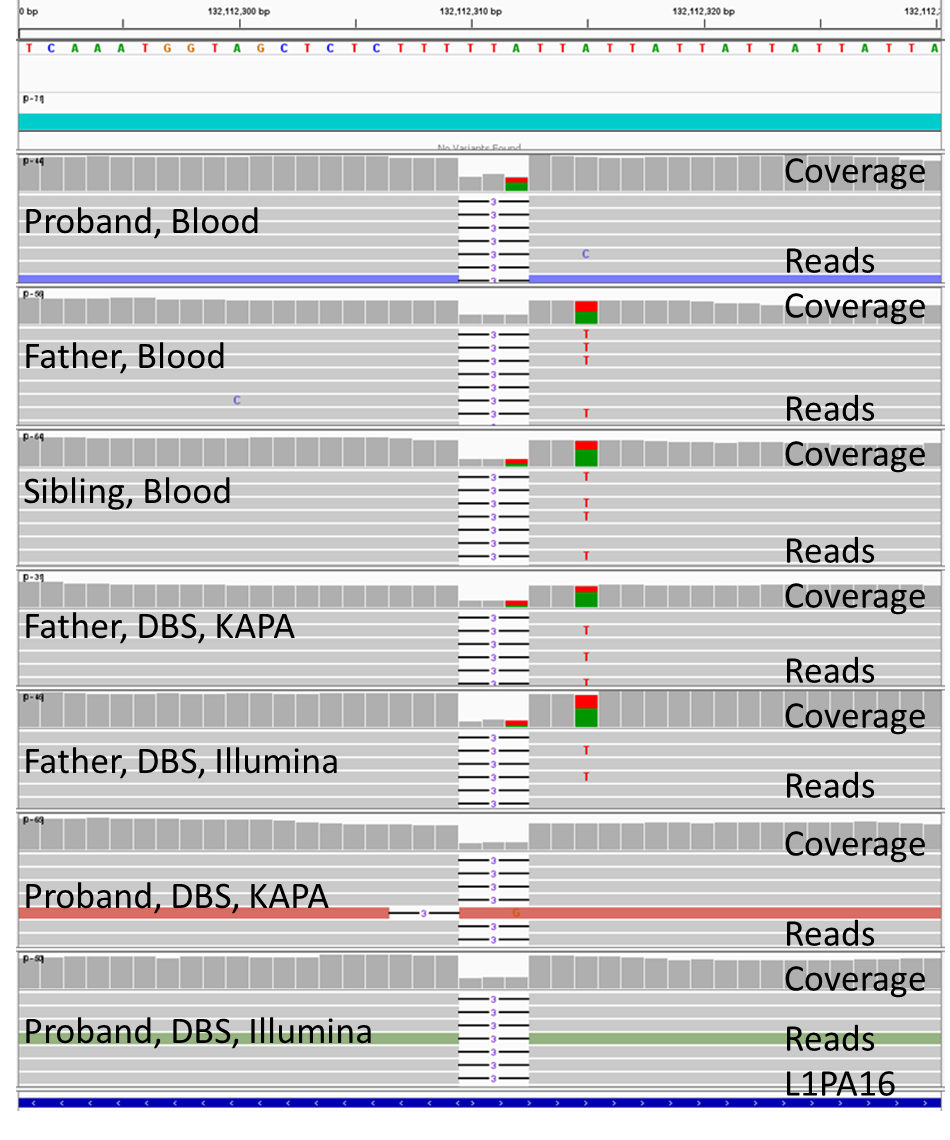

**Figure S7:** Chr 13:64,108,481-64,108,519, a non-coding LINE1 repeat-containing region showing a discrepant 12-nucleotide homozygous or heterozygous deletion based on depth of coverage for the two alleles. Shown, from top to bottom, are the reference nucleotide sequence, average WGS coverage in 200 unrelated subjects, and coverage and representative reads from a proband (blood sample), father (blood sample), sibling (blood sample), father (DBS, KAPA library) father (DBS, Illumina library), proband (DBS, KAPA library), proband (DBS, Illumina library), and RepeatMasker.

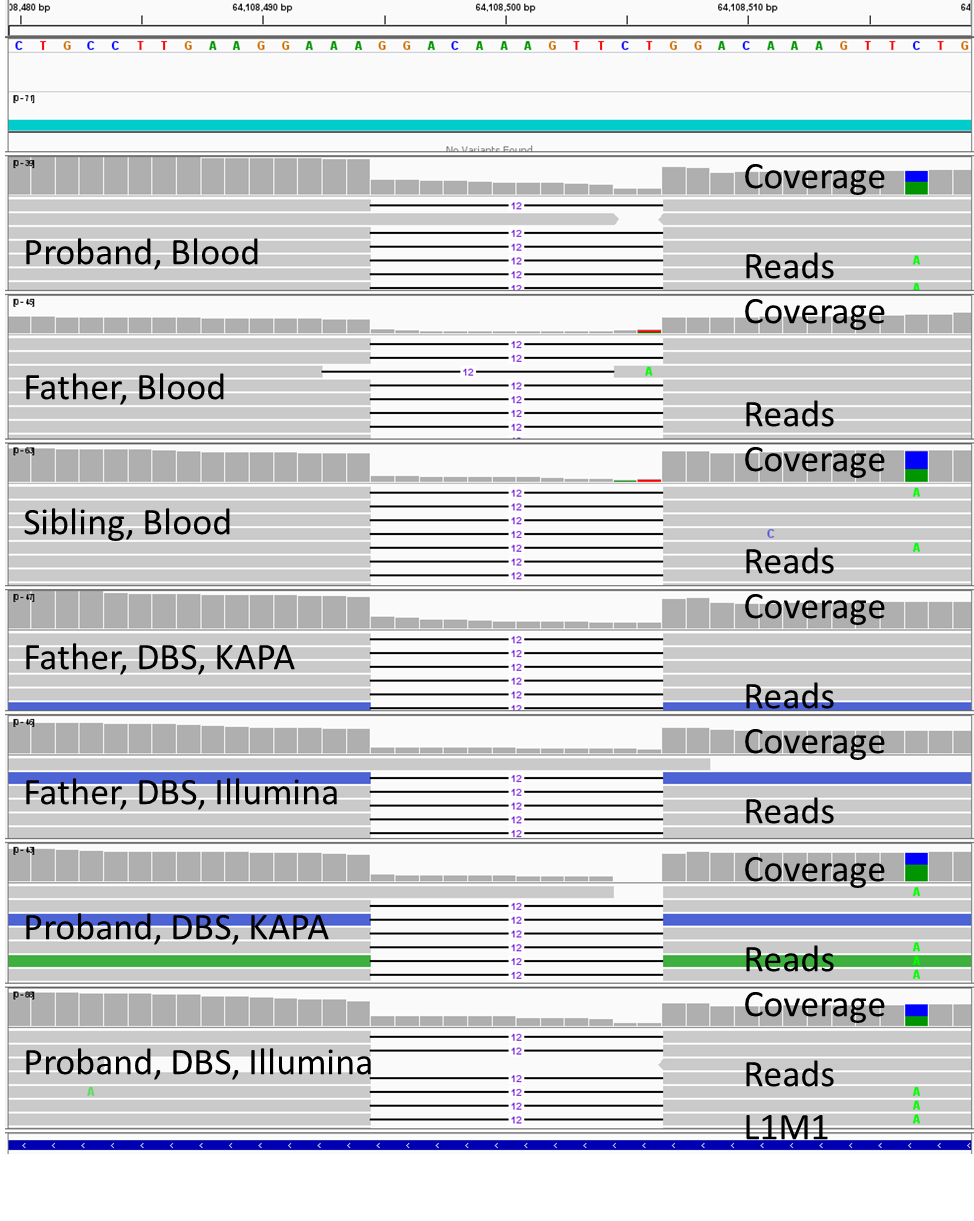

**Figure S8:** Chr 4:171,247,140-171,247,178, a non-coding, AT-rich region, showing a discrepant overlapping homozygous or heterozygous T>A substitution and heterozygous single nucleotide deletion. Shown, from top to bottom, are the reference nucleotide sequence, average WGS coverage in 200 unrelated subjects, and coverage and representative reads from a proband (blood sample), father (blood sample), sibling (blood sample), father (DBS, KAPA library) father (DBS, Illumina library), proband (DBS, KAPA library), proband (DBS, Illumina library), and RepeatMasker.

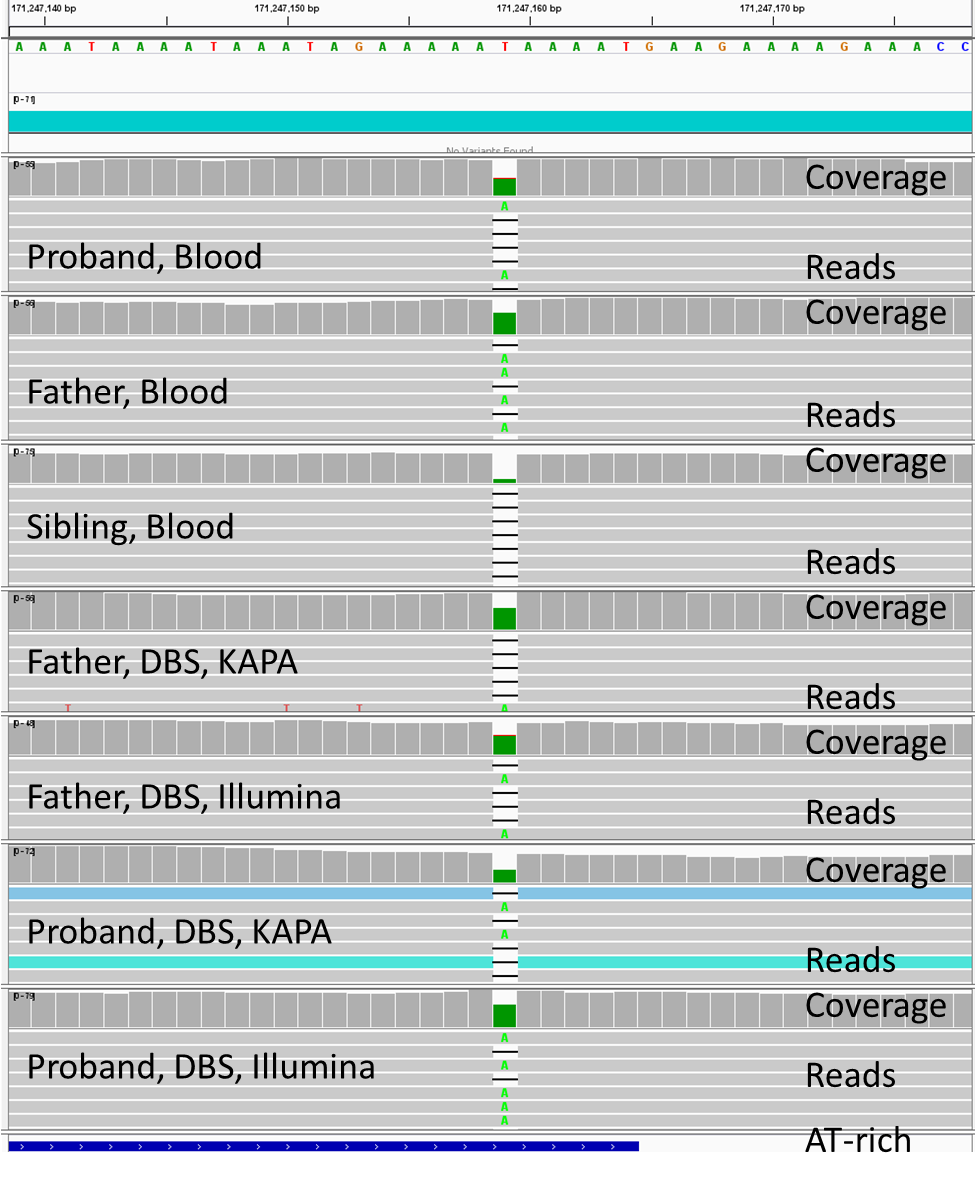

**Figure S9:** Chr 7:67,120,947-67,121,106, a region containing an MSTB1 long terminal repeat endogenous retrovirus, showing a discrepant homozygous or heterozygous C>A substitution. Shown, from top to bottom, are the reference nucleotide sequence, average WGS coverage in 200 unrelated subjects, and coverage and representative reads from a proband (blood sample), father (blood sample), sibling (blood sample), father (DBS, KAPA library) father (DBS, Illumina library), proband (DBS, KAPA library), proband (DBS, Illumina library), and RepeatMasker.

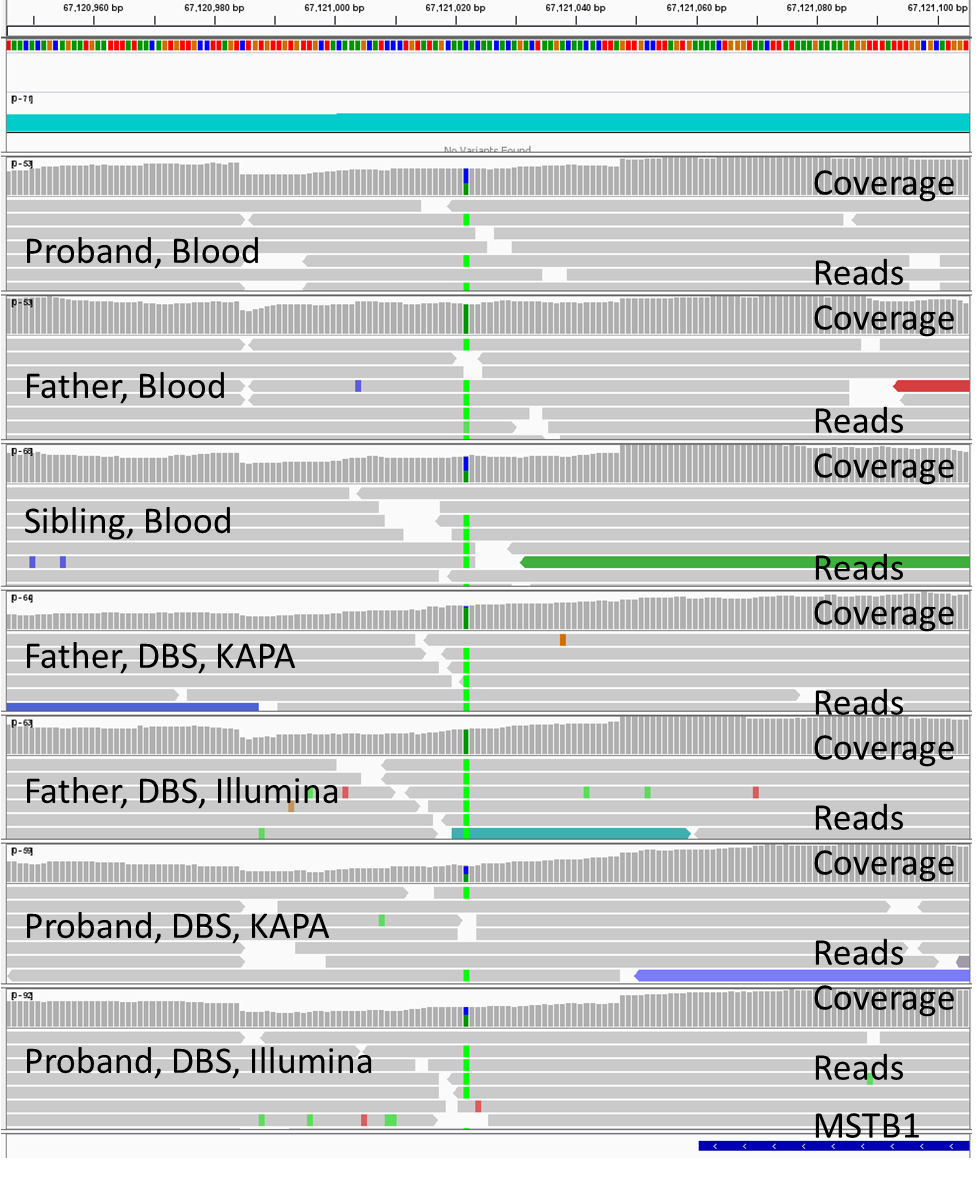

**Fig. S10**: Chr 10:133,159,143-133,159,981, showing a non-coding region that contains a discrepant single versus two nucleotide deletion in a 9 nucleotide guanine homopolymer. Shown, from top to bottom, are the reference nucleotide sequence, average WGS coverage in 200 unrelated subjects, and coverage and representative reads from a proband (blood sample), father (blood sample), sibling (blood sample), father (DBS, KAPA library) father (DBS, Illumina library), proband (DBS, KAPA library), proband (DBS, Illumina library), and RepeatMasker.
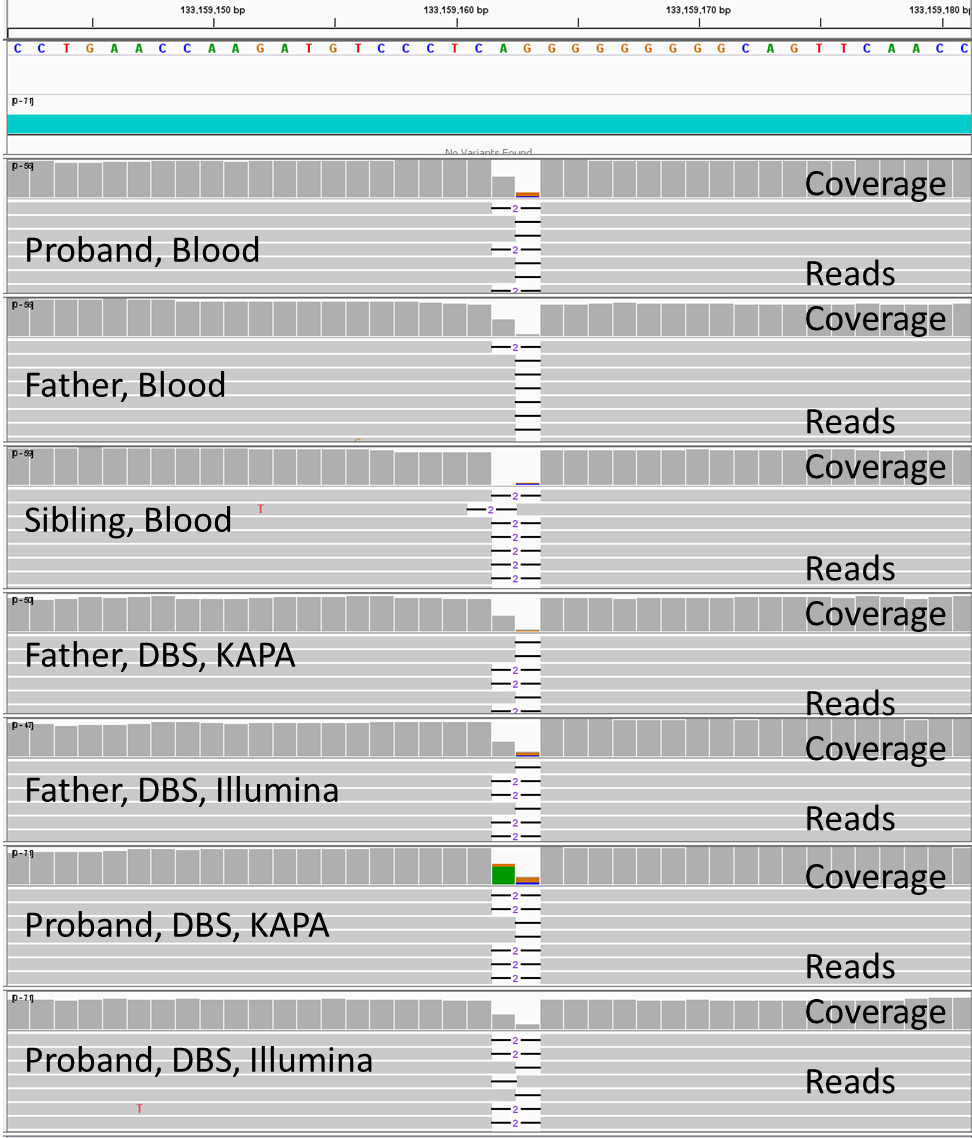

**Fig S11:** Chr 11: 110,204,648-110,204,686 showing a non-coding region that contains an Alu repetitive element, a heterozygous 5 nucleotide deletion within a polythymidine tract, and a discordant, overlapping, heterozygous single nucleotide insertion.

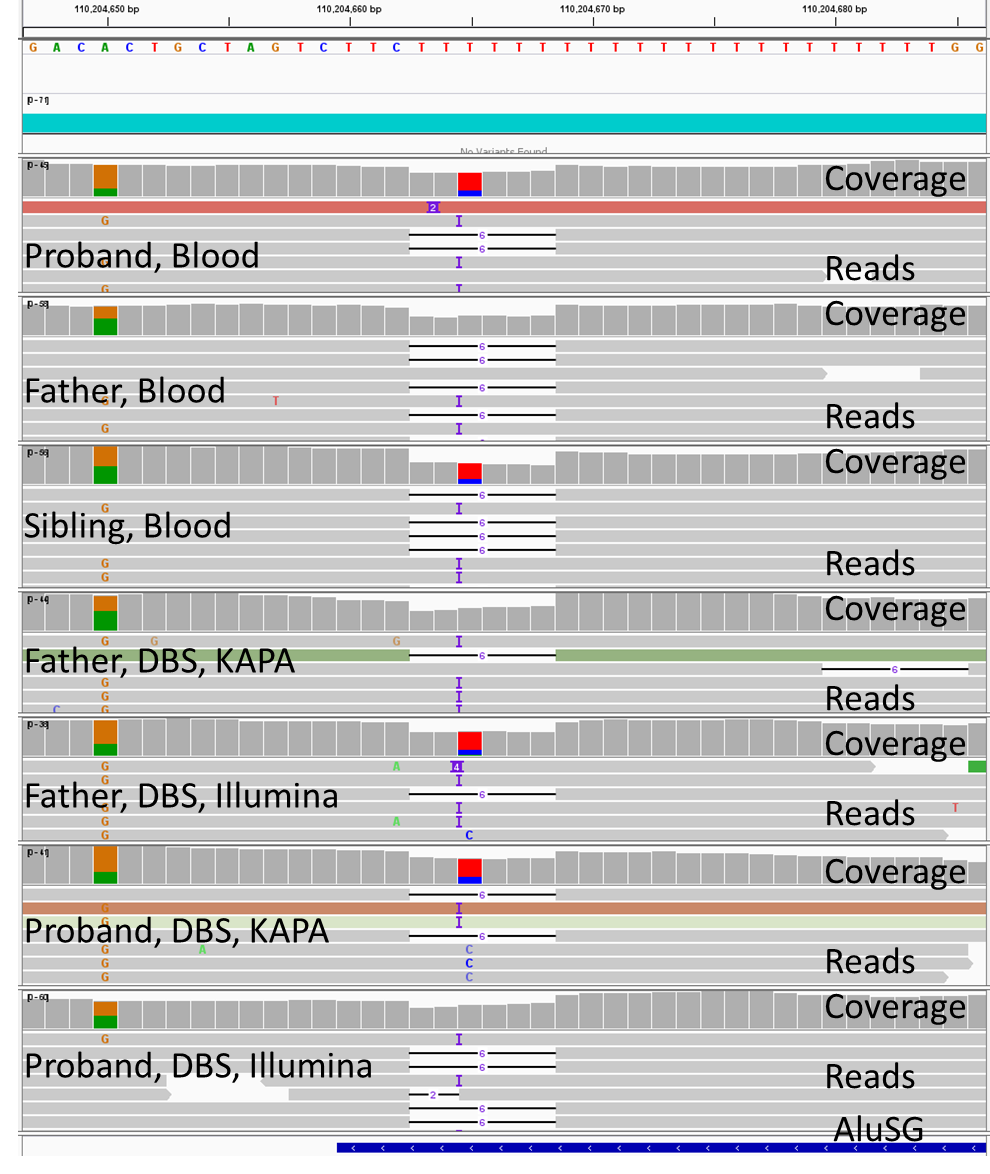

**Fig. S12:** Chr 4:67,030,778-67,030,816, a non-coding polythymidine tract within a MER repetitive element showing a discordant heterozygous A>T substitution overlapping a thymidine mononucleotide deletion.

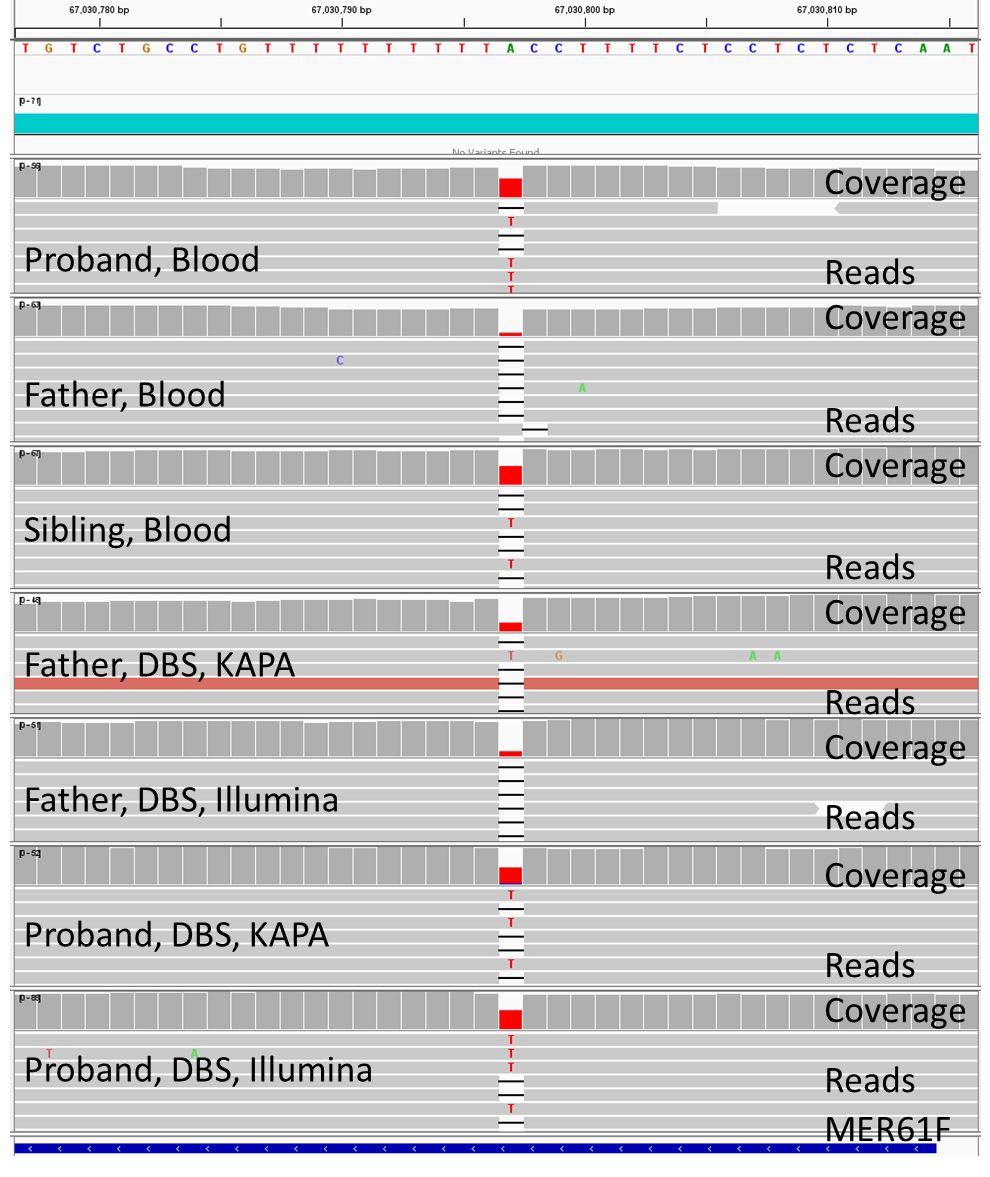

**Fig. S13:** Chr 2: 213,184,938-213,184,976 showing an intron of *ERBB4* containing 4 variants within a polythymidine tract. They are a heterozygous haplotype comprising a single nucleotide insertion and an A>T substitution, a single nucleotide deletion overlapping the A>T substitution (which was called homozygous or heterozygous in different sample types), and a haplotype comprising the A>T substitution (without the insertion) and a C>T substitution.

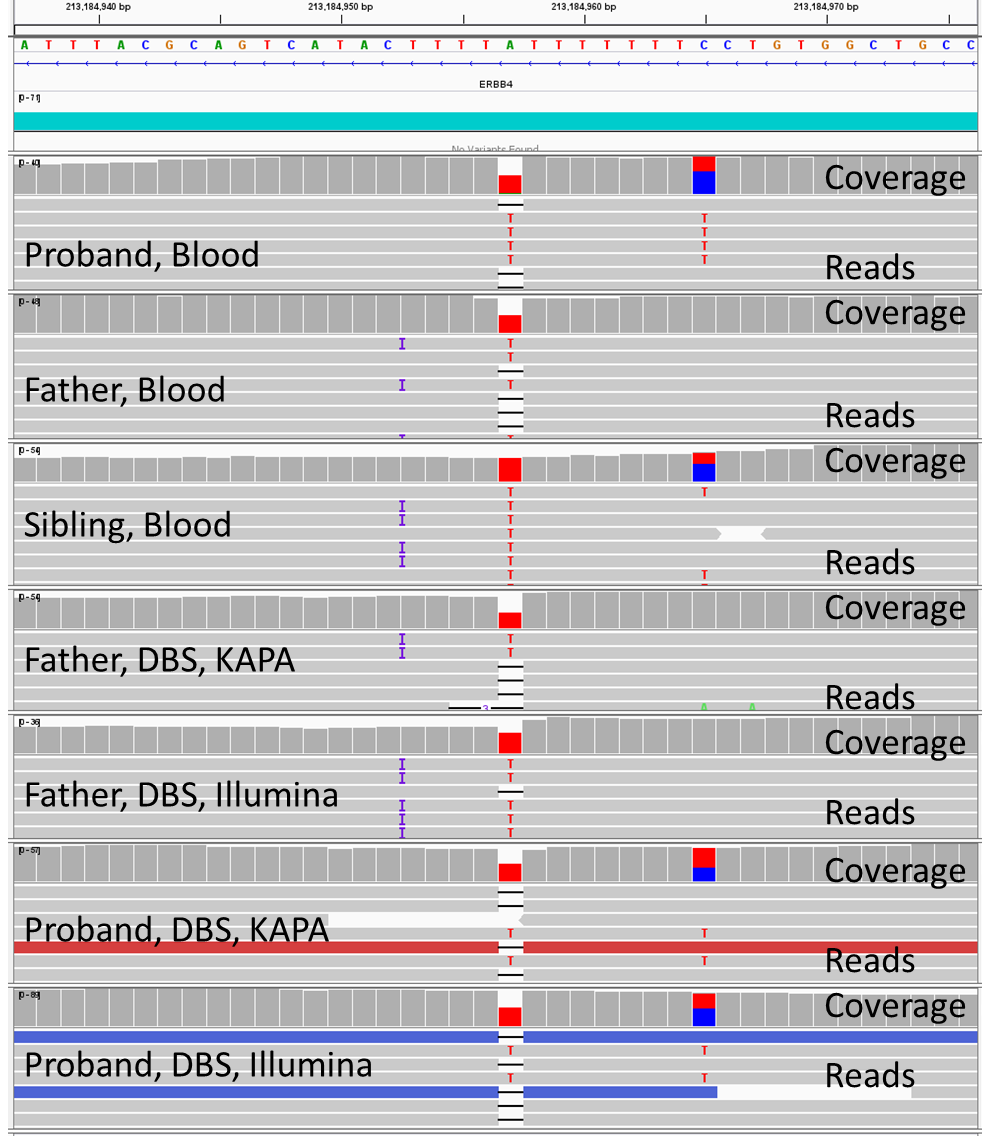

**Fig. S14:** Chr. 14:78,838,500-78,838,538 showing a non-coding pentathymidine tract adjacent to a 9 adenine homopolymer containing a discrepant homozygous or heterozygous thymidine deletion and T>A substitution together with an overlapping heterozygous TT>AA substitution.

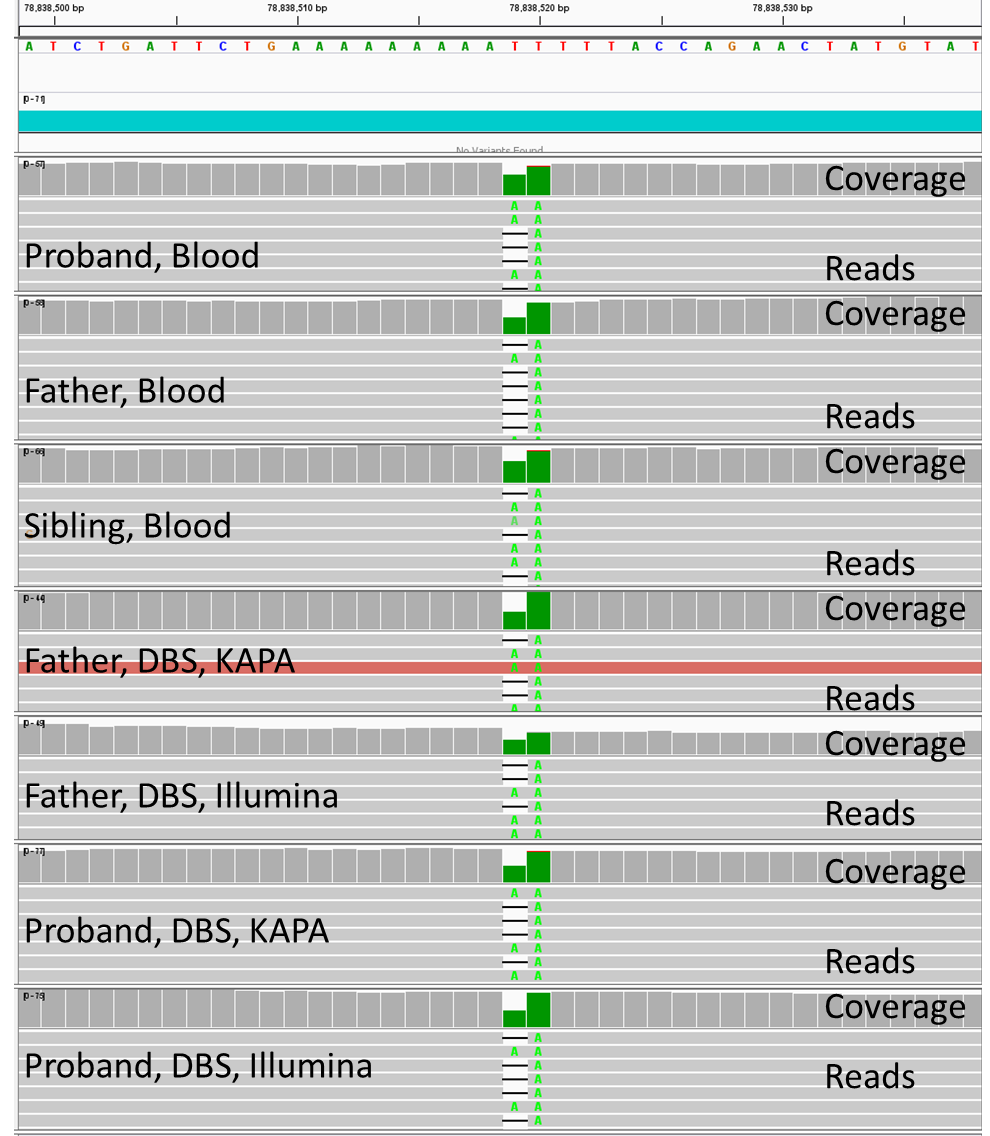

**Fig. S15:** Chr 11:107,991,604-107,991,642 showing a non-coding region with a FLAM element flanked by a homoadenine tract with a discrepant heterozygous versus homozygous A>G substitution and 1-base deletion haplotype in the proband, and an overlapping, discordant, heterozygous versus homozygous A>G substitution in the father.

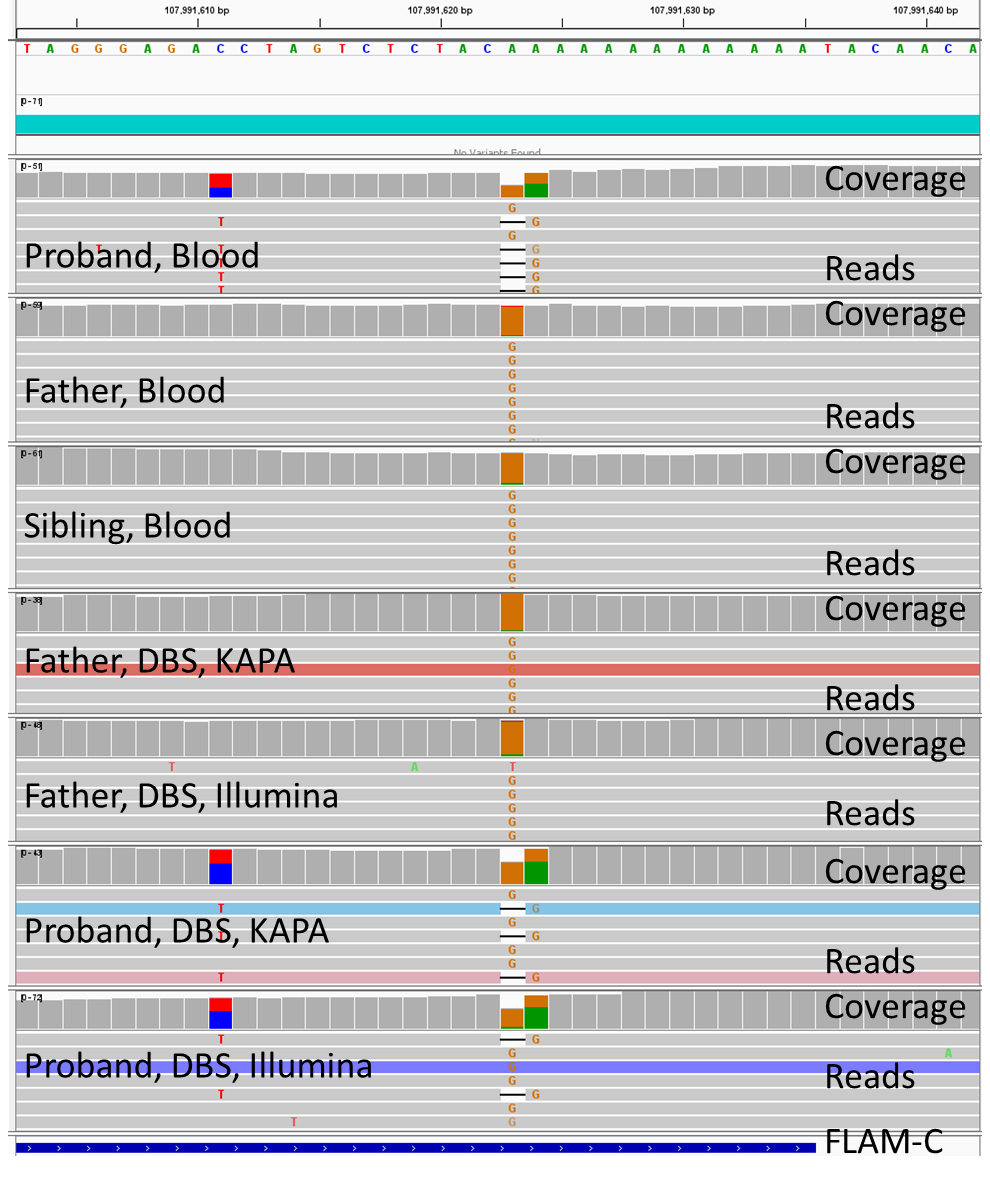

**Fig. S16:** Chr 7:3,185,475-3,185,513, a non-coding region containing an Alu repeat with a discordant heterozygous versus homozygous single nucleotide deletion adjacent to a heterozygous versus homozygous C>T substitution.

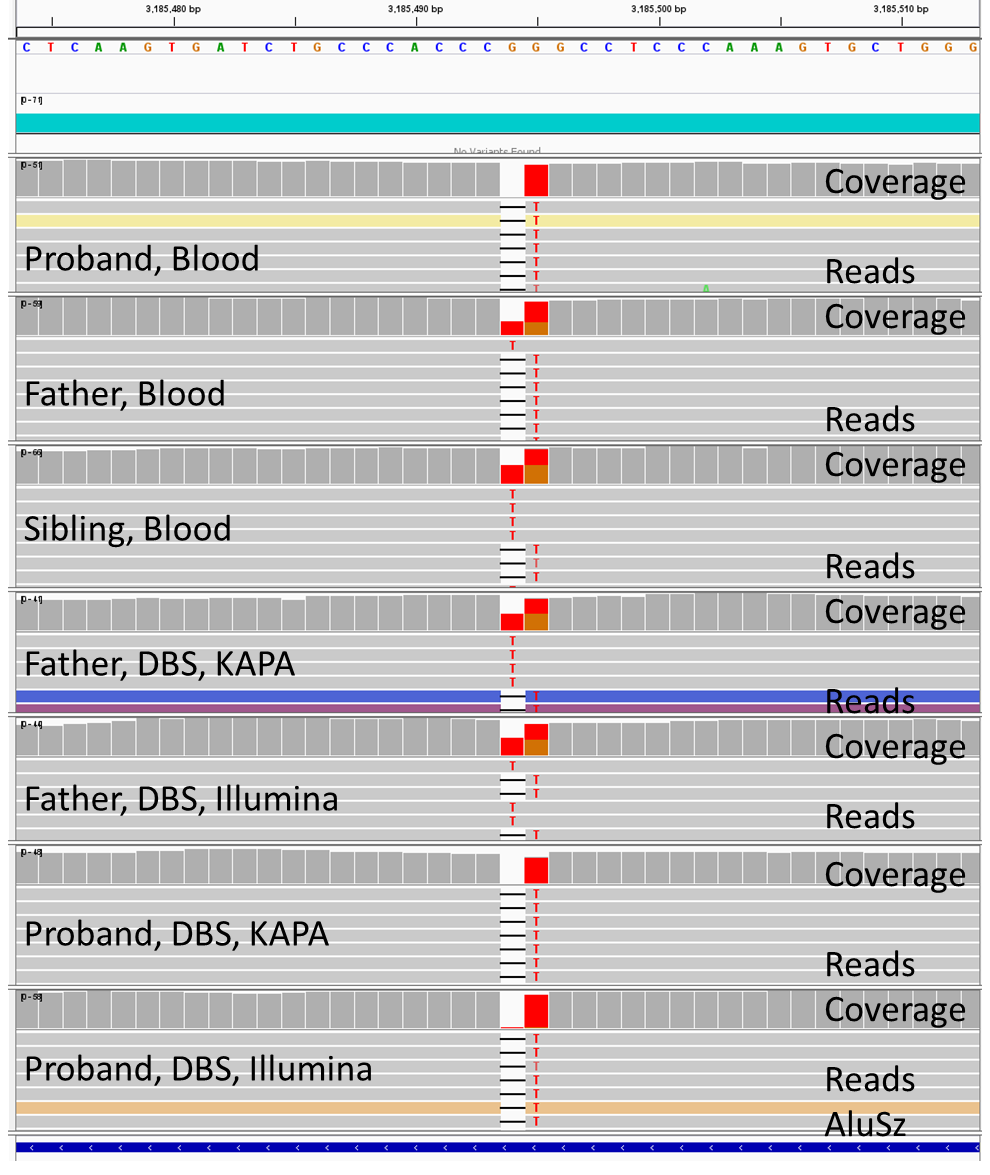

**SUPPLEMENTARY METHODS**

**Simulated DBS specimens using EDTA whole blood**

DBS specimens were collected with Whatman NUCLEIC-CARD^TM^ matrix (ThermoFisher, Catalog #: 4473975) and 903 Protein Saver Cards (GE Healthcare, Catalog #:10534612). Overall, two to ten spots made for each Whatman FTA card or GE Protein Saver card. Preliminary data showed similar final yield of gDNA from spotting 80 µl and 40 µl of blood per disk. Therefore, each spot was made using an equal amount of 40 µl EDTA whole blood to ensure DBS disc size uniformity. The DBS cards were kept at room temperature for at least 2 hours before they dried. Dried cards were stored inside a desiccator at room temperature for later use. The age of DBS from the time it was made to the time when the gDNA was isolated ranged from 1 day to 366 days. Average age of DBS at time of gDNA extraction was 117 days.

**Genomic DNA manual isolation and quality control (QC)**

Each DBS disc for both cohorts was visually examined prior to isolation to ensure no damage had occurred and that there was full absorption on the paper. For cohort 1, two different lysis protocols were used for the four tests performed. Lysis protocol 1 was performed as following: six 3x3 mm punches from a DBS specimen were manually collected in a 1.5 mL Eppendorf tube and mixed with 2 µl proteinase K, 30 µl lysis buffer (Illumina catalog # 2018706 or QIAgen catalog #19075), and 268 µl nuclease free water. The sample tube then was incubated at 56°C for 15 minutes in a thermomixer set at 1000 rpm. Lysis protocol 2 was performed as following: ten 3x3 mm punches were mixed with 4 µl proteinase K, 40 µl lysis buffer and 356 µl nuclease free water. The sample tube was then incubated at 56°C for 60 minutes in a thermomixer set at 2000 rpm. The titration experiment applied different amounts of input at 1, 3, 5, 7 and 10 punches per extraction. For cohort 2, lysis protocol 1 was used for all samples.

For both cohorts, after incubation, the punches/reagents mixture was briefly spun down and the supernatant was carefully transferred into a new Eppendorf tube without disturbing the DBS residuals. 135 µl (for Lysis Protocol 1) or 175 µl (for Lysis Protocol 2) of well-mixed, room temperature normalized KAPA pure beads (Roche/KAPA Biosystems, Catalog #: KK8002) was added into the tube and the solution was mixed by rotating the tube on a rotator (or equivalent) for 15 minutes at room temperature. The sample tube then was placed on a magnet bar (or equivalent) for 5 minutes, the supernatant was discarded and the pellet was washed twice with 500 µl 80% ethanol. The sample/pure beads were air-dried for a few minutes at room temperature before genomic DNA was eluted using 20 to 40 µl elution buffer (10mM Tris-HCl, pH 8 to 8.5). Genomic DNA (gDNA) then was quantified and qualified using Picogreen assay and Nanodrop A260/A280 assays, by following manufacturer’s protocols^21, 22^. Electrophoresis using 0.8% E-gel (ThermoFisher, catalog # A25798) was performed for a subset of selected gDNA samples to evaluate the integrity of the extracted gDNA. The manual isolation of the lysis 1 and lysis 2 protocol took approximately about 60 minutes and 100 minutes respectively.

**Manual sequencing library Preparation**

For cohort 1, two library construction procedures were tested and are detailed below: 1) Illumina PCR-free Tagmentation library kits (Illumina) and 2) KAPA Hyperplus PCR-free library kits (KAPA). The KAPA Hyperplus PCR-free method was repeated using the subset of DBS aimed at assessing reproducibility of the procedures. For cohort 2, sequencing libraries were prepared using DNA PCR-free Tagmentation library prep kit (Illumina) according to the manufacturer’s instructions. Both the Illumina and KAPA method for PCR-free library took approximately 3 hours. The two methods are described below.

1. **PCR-free library construction using the Illumina PCR-free Tagmentation kits* (The kits were provided by the Illumina for assay development, Cat#: 20041855) and QC**

An average of 286 ng gDNA in 10 mM tris-HCl (pH 8 or 8.5) solution was isolated from each DBS and incubated with 10 µl tagmentation buffer and 10 µl beads-linked transposome at 41°C for 5 minutes. 10 µl stop tagmentation buffer was added and well mixed, then incubated at room temperature for 5 minutes to stop the tagmentation reaction. The sample mixture was placed on a magnet bar or plate until the solution was clear, then about 60 µl supernatant was discarded and 150 µl tagmentation wash buffer was added while the sample was kept on the magnet bar or plate. The 150 µl tagmentation wash buffer was then removed. For the ligation step, 45 µl extension ligation mix and 5 µl index adaptor were both added, and the sample mixture was incubated at 37°C for 5 minutes and 50°C for 5 minutes. Using 75 µl tagmentation wash buffer, the products were washed while keeping the sample mixture on the magnet bar or plate. 75 µl tagmentation wash buffer was then discarded and 47 µl sodium hydroxide (2N) was added into the sample, and incubated at room temperature for 5 minutes. The sample mixture was placed on the magnet bar or plate again and the supernatant was removed. Finally, beads-based double size selection was performed to ensure the fragment size of each sample was within from 450 to 650 bp, following manufacturer protocol^30^. The concentration of ligated fragments in a library was quantified with KAPA Library Quantification Kits for Illumina® platforms (Roche/KAPA Biosystems, Catalog#: KK4824) on Roche LightCycler® 480 Instrument (Roche, Basel, Switzerland). The libraries with concentration more than 3 nM were passed and sequenced on Illumina Novaseq 6000 S4 Flow Cell. Automated Tagmentation library construction is under development at the time of writing.

2. **PCR-free library construction using the Roche KAPA HyperPlus kits (Cat#: KK8515) and QC**

PCR-free library construction was prepared with an average 400 ng of extracted gDNA in a PCR plate. For enzymatic fragmentation, the gDNA was normalized to 30 µl in the suspension buffer. 20 µl fragmentation mixture containing 5 µl diluted conditioning buffer (13.5 µl original conditioning buffer in 86.5 µl nuclease free water), 5 µl fragmentation buffer and 10 µl fragment enzyme were added to each sample well and the plate was incubated at 37°C for 8 minutes. After incubation, a pre-made 10 µl End-Repair (ER) and A-tailing (AT) mixture (7 µl End-Repair/A-tailing buffer, 3 µl enzyme) was immediately added into each sample well and the plate was incubated at 65°C for 30 minutes. In the ligation step, 48 µl ligation master mix (30 µl ligation buffer, 10 µl DNA ligase, 8 µl PCR-grade water) and 2 µl dual index adapter (IDT, San Diego, CA) were added to each well containing sample/ER/AT mix, and the plate was incubated at 20°C for 30 minutes in a thermocycler. Samples were cleaned up using 1x SPRI (Solid Para-magnet Reversible Immobility) beads and 80% ethanol. Finally, beads-based double size selection was performed to ensure the fragment size of each sample was within from 450 to 650 bp, following manufacturer protocol^31^. The fragment size of a DNA library sample was measured using Agilent DNA High Sensitivity NGS Fragment Analysis Kit (Agilent, Catalog#: DNF-474-0500) to ensure it was between 300 bp to 600 bp. Concentration of a library was also quantified using KAPA Library Quantification Kits for Illumina Platforms described aforementioned section. The libraries with concentration more than 3 nM and acceptable fragment size were passed and sequenced on Illumina Novaseq 6000 S1 or S2 Flow Cell. Fully automated KAPA HyperPlus library preparation has been validated for clinical samples, although it was not used here.

**Whole genome sequencing**

For cohort 1, two and half libraries (sequencing on a S1 FC), five to six libraries (sequencing on a S2 FC), or twenty four libraries (sequencing on a S4 FC) were pooled with an equal molarity for a final loading concentration between 400 pM to 450 pM. Cohort 2 was run on either the S1 flow cell or the SP flow cell. The pooled libraries were denatured with 0.2 N sodium hydroxyl for 8 minutes. 400 mM Tris-HCl (pH 8.5) was then added to terminate denaturing reactions. The flow cells were loaded on the Illumina Novaseq 6000 with a read length of 2x101 or 2x 151 (IDT dual indexing, Cat#:263582653) for cohort 1, or 2x250 or 2x100 for cohort 2. Whole genome sequencing with high quality sequencing raw data targeted was a FC with Q30≥ 80% and 120 Gb or greater per WGS. If the first sequencing attempt generated less than 120 Gb per sample, an additional top-off sequencing run was performed.
